# The impact of technology systems and professional support in digital mental health interventions: a secondary meta-analysis

**DOI:** 10.1101/2021.04.12.21255333

**Authors:** Maxime Sasseville, Annie LeBlanc, Jack Tchuente, Mylène Boucher, Michèle Dugas, Mbemba Gisèle, Romina Barony, Maud-Christine Chouinard, Marianne Beaulieu, Nicolas Beaudet, Becky Skidmore, Pascale Cholette, Christine Aspiros, Alain Larouche, Guylaine Chabot, Marie-Pierre Gagnon

**Affiliations:** Université du Québec à Chicoutimi, VITAM Research Center on Sustainable Health; Université Laval, VITAM Research Center on Sustainable Health; VITAM Research Center on Sustainable Health; Université Laval; Université de Montréal; Université de Sherbrooke, Omnimed; Independent Information Specialist; Centre Intégré Universitaire de Santé et de Services Sociaux de la Capitale Nationale; Patient-Partner; Groupe Concerto

**Keywords:** Digital mental health, Chronic diseases, Meta-analysis, Rapid Review

## Abstract

**Background:** A rapid review of systematic reviews was conducted to assess the effectiveness of digital mental health interventions for people with a chronic disease. Although it provided an overview of the evidence, it offered limited understanding of ethe types of interventions that were the most effective. The aim of this study was to perform a meta-analysis of primary studies identified in this rapid review of systematic reviews by focusing on the needs of knowledge users.

**Methods:** This secondary meta-analysis follows a rapid review of systematic reviews, a virtual workshop with knowledge users to identify research questions and a modified Delphi study to guide research methods. We conducted a secondary analysis of the primary studies identified in the rapid review. Two reviewers independently screened the titles and abstracts and applied inclusion criteria: RCT design using a digital mental health intervention in a population of adults with another chronic condition, published after 2010 in French or English, and including an outcome measurement of anxiety or depression.

**Results:** 708 primary studies were extracted from the systematic reviews and 84 primary studies met the inclusion criteria Digital mental health interventions were significantly more effective than in-person care for both anxiety and depression outcomes. Online messaging was the most effective technology to improve anxiety and depression scores; however, all technology types were effective. Interventions partially supported by healthcare professionals were more effective than self-administered.

**Conclusions:** While our meta-analysis identifies digital intervention’s characteristics that are more effective, all technologies and levels of support can be used considering implementation context and population.

**Review registration:** The protocol for this review is registered in the National Collaborating Centre for Methods and Tools (NCCMT) COVID-19 Rapid Evidence Service (ID 75).

## BACKGROUND

Chronic diseases are the main burden on health systems in developed countries and account for almost 70% of deaths worldwide (1). The majority of people with a chronic disease have more than one concurrent condition and are also at higher risk for developing comorbidities in mental health, including anxiety and depression (2). The prevalence of depression together with another chronic health disease is an important and well-documented phenomenon (3). Studies have also reported a higher prevalence of anxiety disorders in relation to different chronic diseases (4). Increased use of health services is also observed in people living with a chronic disease and with a concomitant depressive disorder (5). Canadians primary healthcare interdisciplinary teams, surveyed during the COVID-19 pandemic, reported a need for broadening services offering to answer the increase in encounters for mental health issues (6). There is an urgent need for more relevant and accurate data on digital interventions in this area to prepare for an increase demand for mental health services.

Practice-based interventions in primary care settings have been shown effective to improve the management of depression in people with chronic diseases (3). In addition, a large number of interventions using digital technologies have been evaluated for the management of depression or anxiety (7, 8), and systematic reviews indicate that they are effective in providing timely and delocalized care in some populations (9). However, it is still unclear what elements or characteristics of digital interventions for mental health are effective (9).

A rapid review provides knowledge users with data that can be readily used to inform healthcare decisions (10). When the topic is broad, it can lead to data that lack precision to fulfil knowledge users’ needs. In an effort to gather data on the effectiveness of digital mental health interventions for people with a chronic disease, a rapid review of systematic reviews was completed (11) and offered only an overview of the problem. The aim of this study was to conduct a secondary meta-analysis of the data collected in this rapid review of systematic reviews by using a knowledge-user panel to guide research methods.

## METHODS

We engaged with a panel of knowledge users (patients, clinicians, decision makers), content experts, review methodologists, and researchers throughout the review process, including question development, literature search, interpretation and writing of results, and dissemination of findings.

### Preliminary research

This secondary analysis is based on the data from a rapid review of systematic reviews (11). We followed guidelines outlined by the Cochrane Handbook chapter regarding Overview of Reviews and the Cochrane Rapid Reviews Methods Group (12, 13). The review identified a large body of evidence (35 systematic reviews) showing that digital mental health interventions were effective and safe for people with chronic diseases and cancer but that the evidence was still lacking for children and youth populations. In order to inform the knowledge users at each step, the first stage and lessons learned while developing the project were published elsewhere (11, 14).

Three research activities followed the rapid review: a virtual workshop with knowledge users to present our preliminary results and gather their suggestions, a modified Delphi study to prioritize the proposed suggestions for the next stage of the review, and a secondary analysis of the primary studies identified through the rapid review. All activities of the study are summarized in Figure 1.

**Figure 1.**
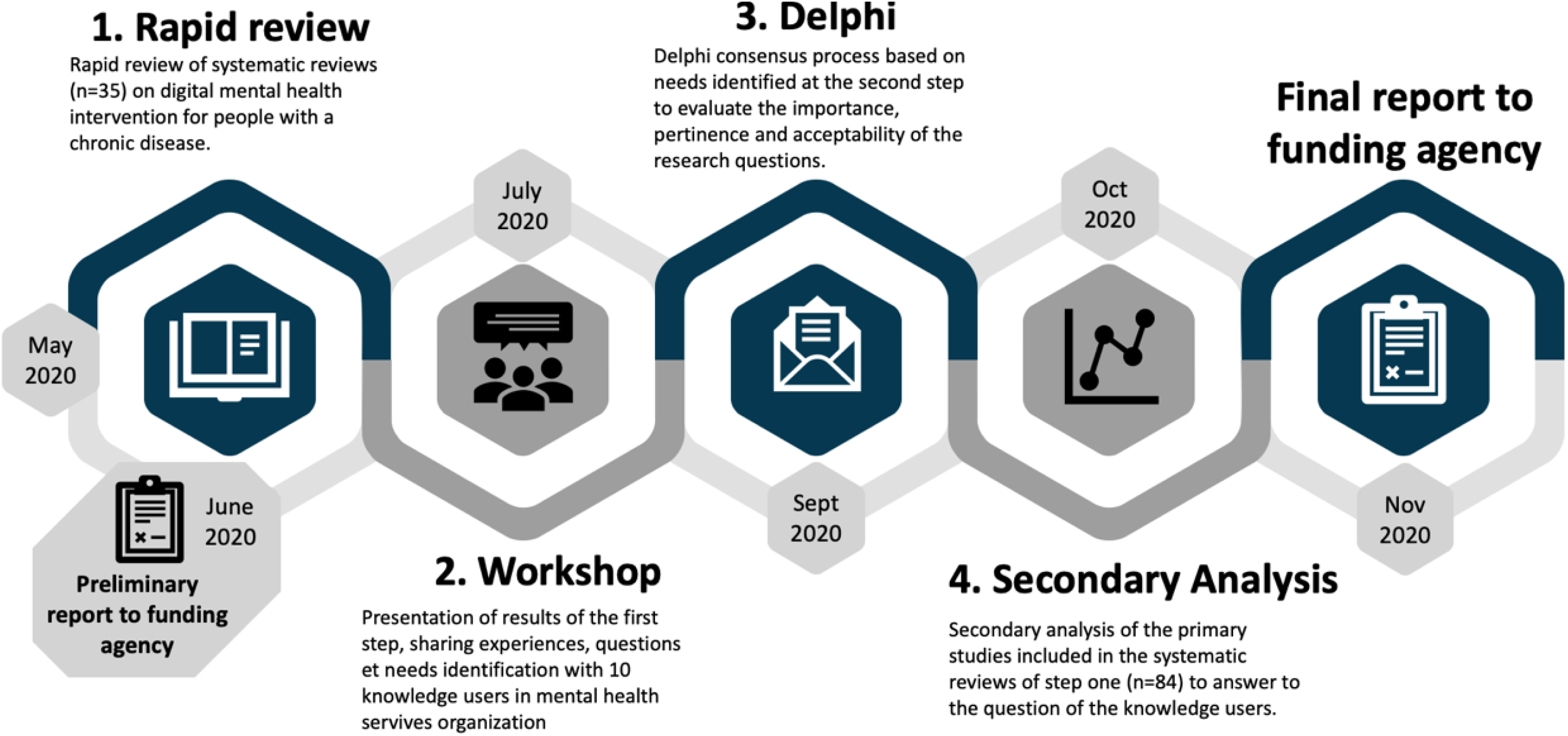
Summary of the activities and timeline

#### Workshop

A total of 10 knowledge users, from provincial (Quebec Ministry of Health and Social Services, Institut national d’excellence en santé et services sociaux), regional (Integrated Health and Social Services Centers) and local organizations were invited to take part in a virtual meeting on July 16^th^ 2020 where the preliminary results from the rapid review were presented and discussed. During the workshop, knowledge users were invited to share their experience regarding digital health interventions for mental health issues, in particular in the context of the COVID-19 pandemic. They were welcome to ask questions and to comment on the findings of the rapid review and its relevance for their practice. They were also encouraged to identify knowledge gaps that could be addressed in the next stage of the review.

#### Delphi Study

Following the workshop with knowledge users, a summary of the main knowledge gaps identified was performed by the research team. We translated these knowledge gaps into nine review questions that were used as the basis for a modified Delphi study. We developed a questionnaire using the RedCap system (15), and sent a personalized invitation to participants in the workshop, inviting them to complete the survey. These knowledge users were also invited to suggest names of potential additional knowledge users who could have an interest in the topic. The questionnaire comprised two sections. First, participants had to rate on a 5-point Likert scale the importance, relevance, and applicability in the context of COVID-19, each of the nine potential review questions. Second, participants were invited to rank each of the nine questions (1 = most important; 9 = less important) according to their preference. A total of 16 knowledge users were invited to take part in a two-round Delphi process in order to identify the key question for the next stage of the knowledge synthesis. All the knowledge users completed the two rounds and all questions reached consensus using median and IQR. The prioritized question after the second round was: “*What types of digital health interventions (according to a recognized categorization) are the most effective for the management of concomitant mental health and chronic disease conditions in adults*?”

#### Secondary Analysis

To provide an answer to knowledge users, we conducted a secondary analysis of the primary studies included in the reviews identified in our previous rapid review, published elsewhere [1]. Based on the input from knowledge users, we added inclusion criteria at this stage. Thus, we included only randomized-controlled trials (RCT) presenting a digital health intervention for the management of concomitant mental health and chronic disease conditions in adults, published during the last 10 years (since 2010) in French or English, and including an outcome measurement of anxiety and/or depression. We focussed on anxiety and depression because it was the most prevalent outcomes to answer the question of knowledge users.

All the primary studies referenced in the 35 systematic reviews were extracted and included for the citation screening. Six reviewers individually performed screenings for titles, abstracts and then full text using pilot-tested standardized forms (25 citations for the first level of screening). All citations were reviewed by two reviewers independently at the first level of screening. A standardized extraction form was developed that included study characteristics (e.g., authors, country, design), intervention characteristics (e.g., type of digital intervention), and outcomes reported (measurement tools used, means, standard deviation and time of measurement). Data were extracted by four research associates, and a senior investigator (MPG) completed a quality appraisal of all extracted data. Information related to the study characteristics (first author, date of publication, country, population, health condition, type of intervention, outcomes measured, means, standard deviations, author’s conclusions) were extracted directly in the DistillerSR tool (16).

As knowledge users were interested in obtaining evidence on specific types of digital health interventions for concomitant mental health and chronic health conditions, we looked for existing classifications of digital health tools to sort interventions. We consulted the WHO *Classification of digital health interventions (17)* and previous reviews on digital mental health solutions (18, 19). Given the limitations of existing categorizations, we used our own system which considered the technology system, the synchronous or asynchronous nature of the intervention, and the level of professional support (self-administration, partially guided, guided). The classification used ten technology systems categories (not exclusive): Internet or Website, Computer software, Mobile application, Electronic messaging (email, SMS), Electronic health record, Telehealth (telemedicine, telepsychiatry), Virtual reality/ augmented reality, Robot, Connected devices and Other system.

We completed a meta-analysis of the standardized means difference (SMD) with an analysis of heterogeneity (x^2^ and I^2^) for the two outcomes of interest. We used Cohen D, fixed effects meta-analysis and the R software for data analysis.

The Cochrane Risk of Bias for Intervention Studies (ROBIS) tool was completed by two investigators (MS, MPG) to assess the probability of bias in the included studies. Five types of biases were considered: 1) Risk of bias arising from the randomization process; 2) Risk of bias due to deviations from the intended interventions; 3) Missing outcome data; 4) Risk of bias in the measurement of the outcomes; and 5) Risk of bias in the detection of the reported results. An overall risk of bias was also assessed.

We report our results based on the Preferred Reporting Items for Systematic Reviews and Meta-Analyses (PRISMA) Statement (20).

## RESULT

### Characteristics of Included studies

The flow diagram of studies included in the secondary analyses is presented in Figure 2. All individual primary studies included in the systematic reviews of the rapid review were considered.

**Figure 2.**
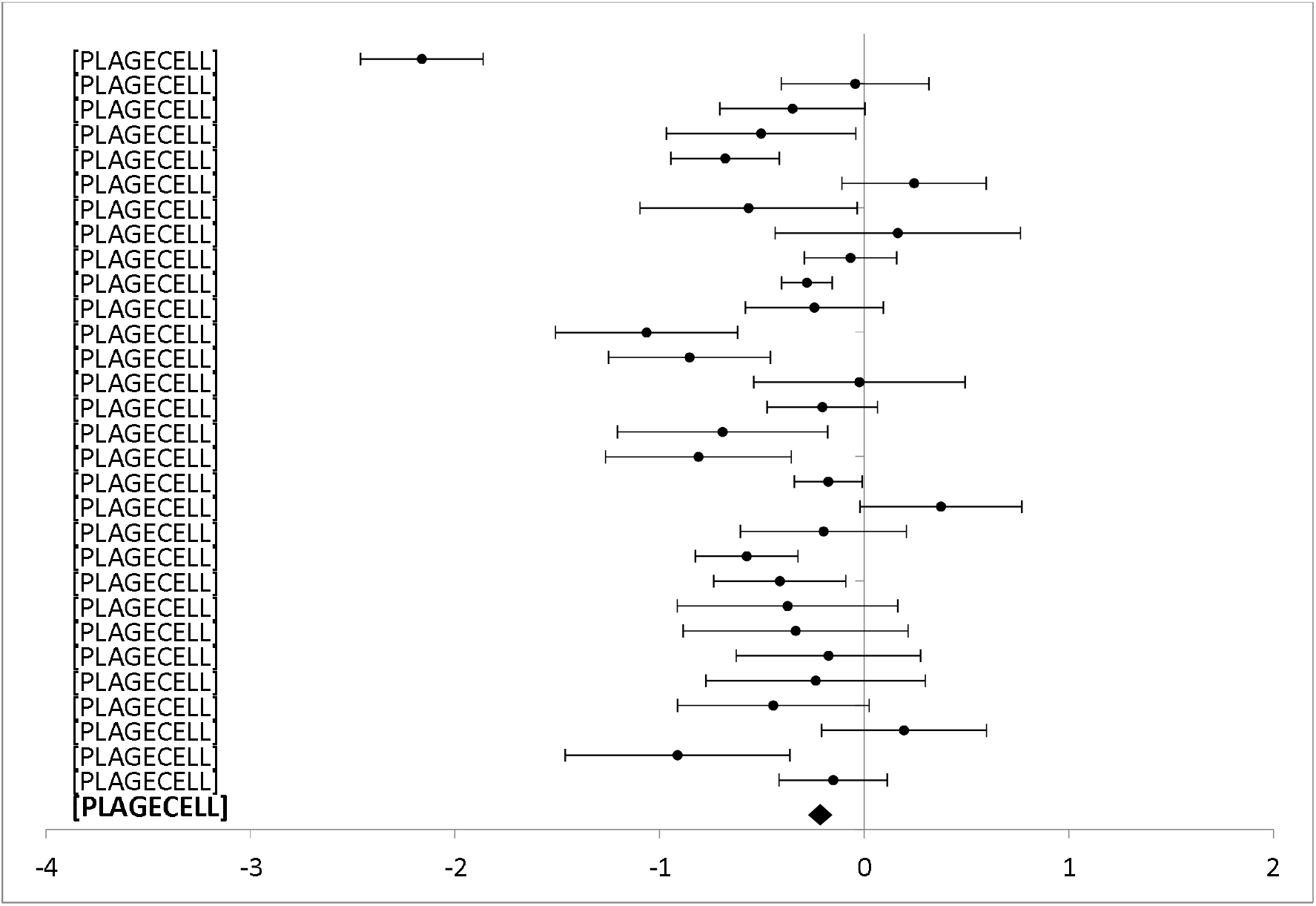
Forest plot: Subgroup analysis of self-directed interventions for any digital intervention vs. usual care or another digital intervention to manage anxiety in people with any concomitant chronic condition.

A total of 708 primary studies were identified from the systematic reviews included in the rapid review. A total of 429 primary studies were excluded at title & abstract screening stage (duplicates: 91; publication year or study design: 338). Following screening of full text, we excluded 195 records with reasons (Population: 40; Intervention: 74; Outcomes: 67; Language: 2), resulting in a total of 84 primary studies included in the secondary meta-analysis.

### Characteristics of Included Studies

We included 84 primary studies, published between 2010 and 2019. Those studies comprised a total sample of 11037 participants. Countries where the studies were conducted included: Sweden (23), the United States (18), Australia (14), Netherlands (9), United Kingdom (7), Germany (6), Switzerland (2), Norway (1), Canada (1), Jordan (1), New Zealand (1) and South Korea (1).

Most studies described interventions performed in the community (33%) and targeted a mixed gender adult population (91%). All studies evaluated digital interventions to manage and treat mental health issues, and a majority (80%) were based on cognitive behavioural therapy (CBT). Most studies compared digital health interventions to usual care (76%), although some studies compared two or more digital interventions (24%). The complete description of included studies is presented in Table 1.

**TABLE 1.**
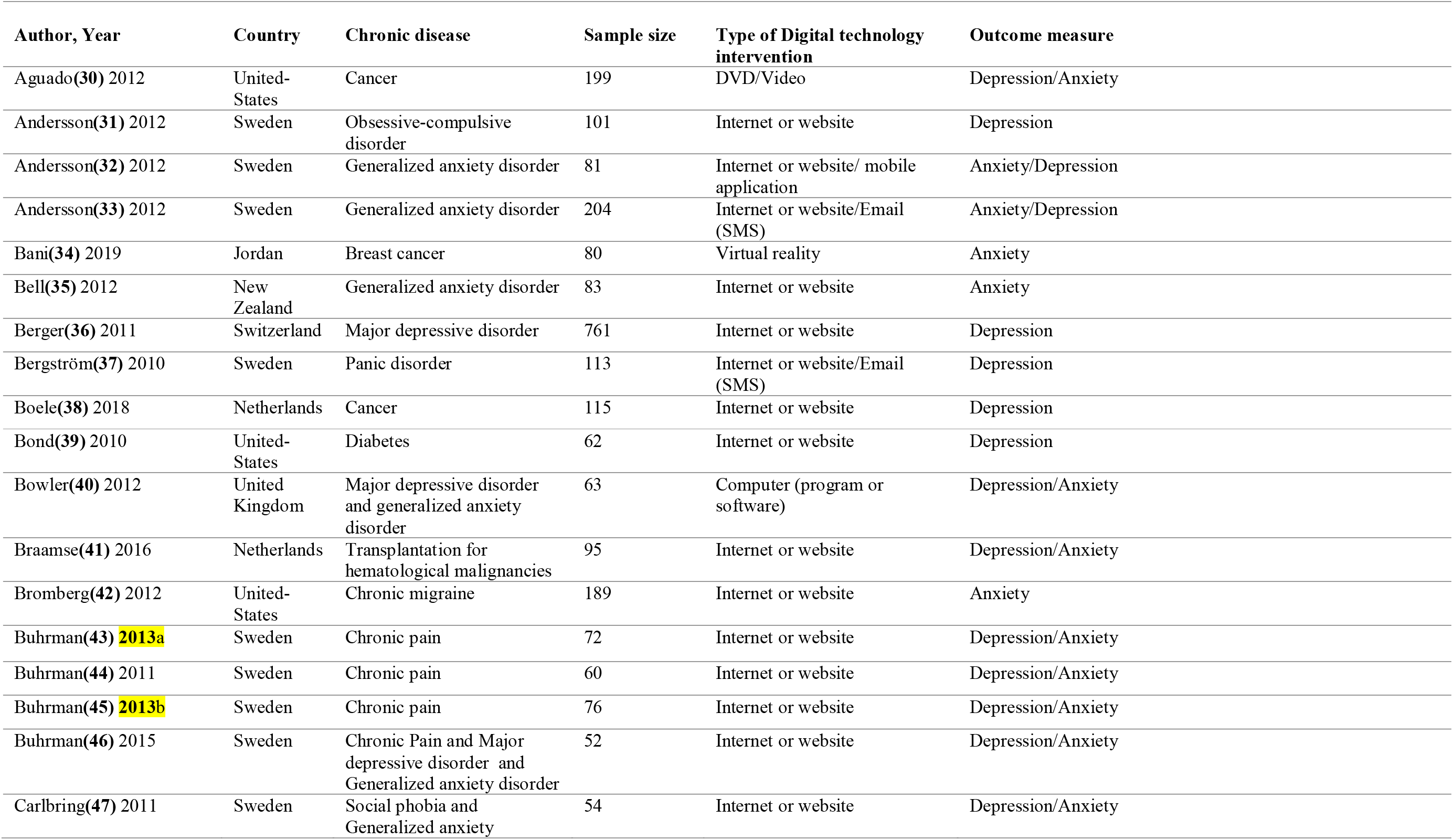
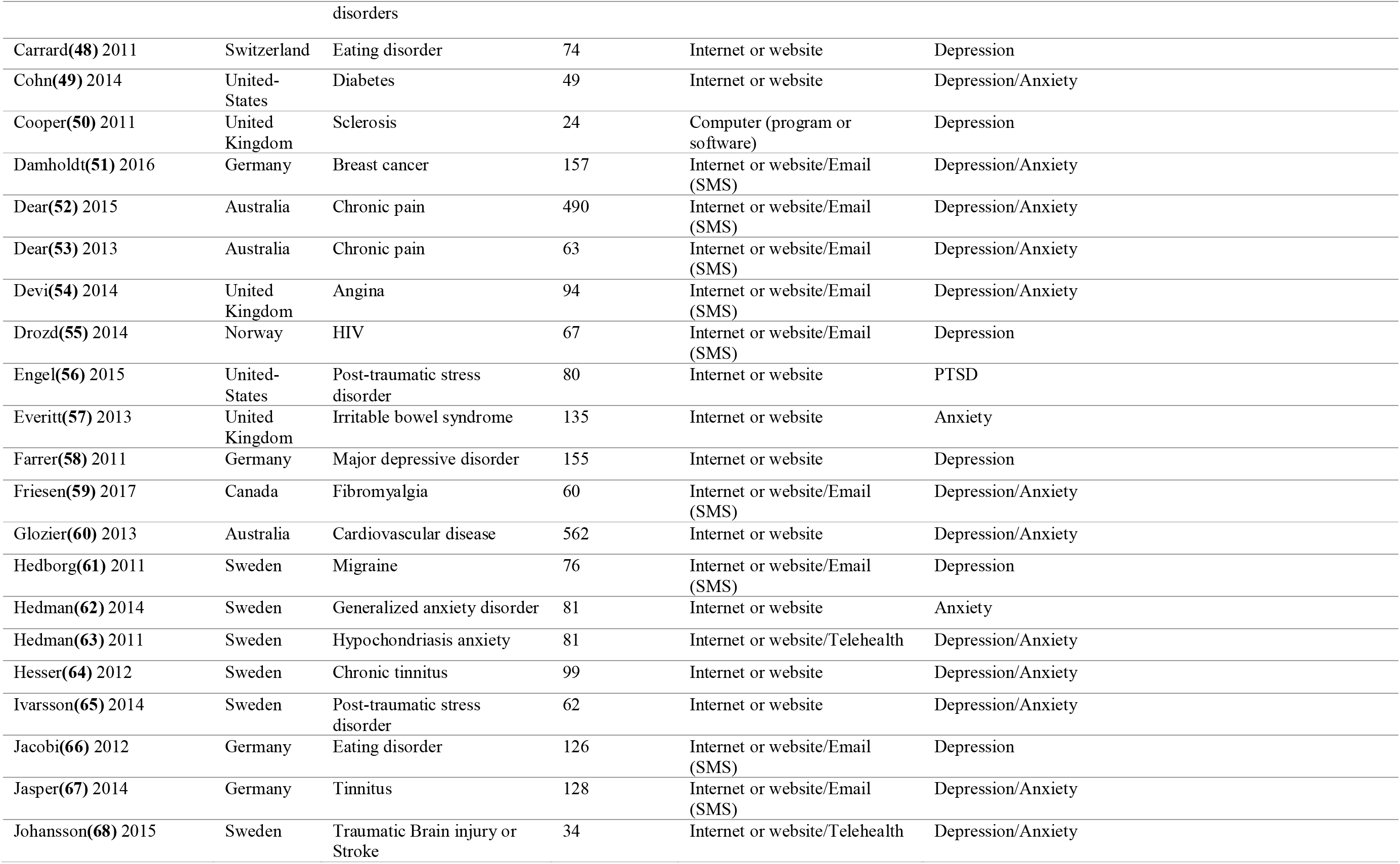
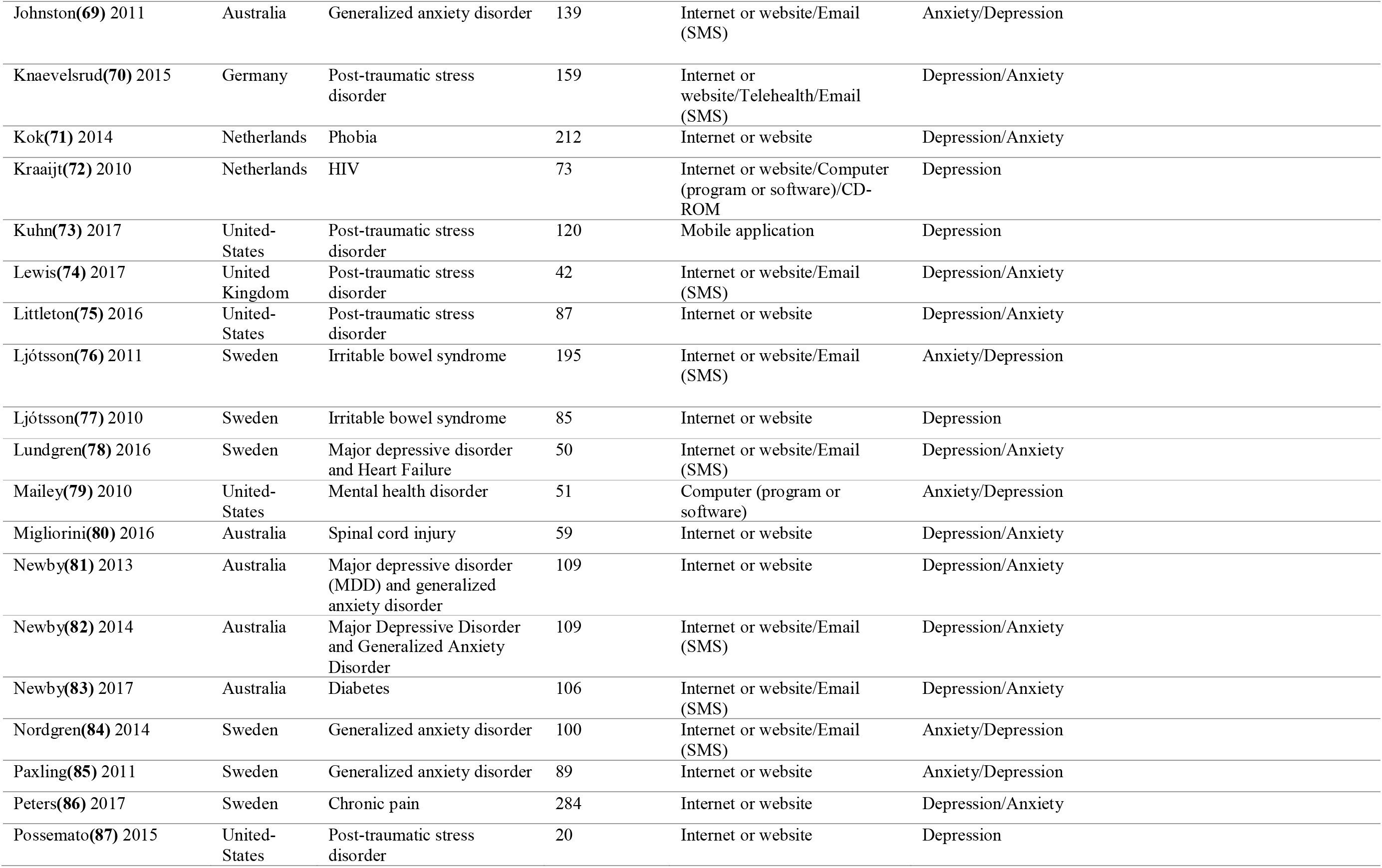
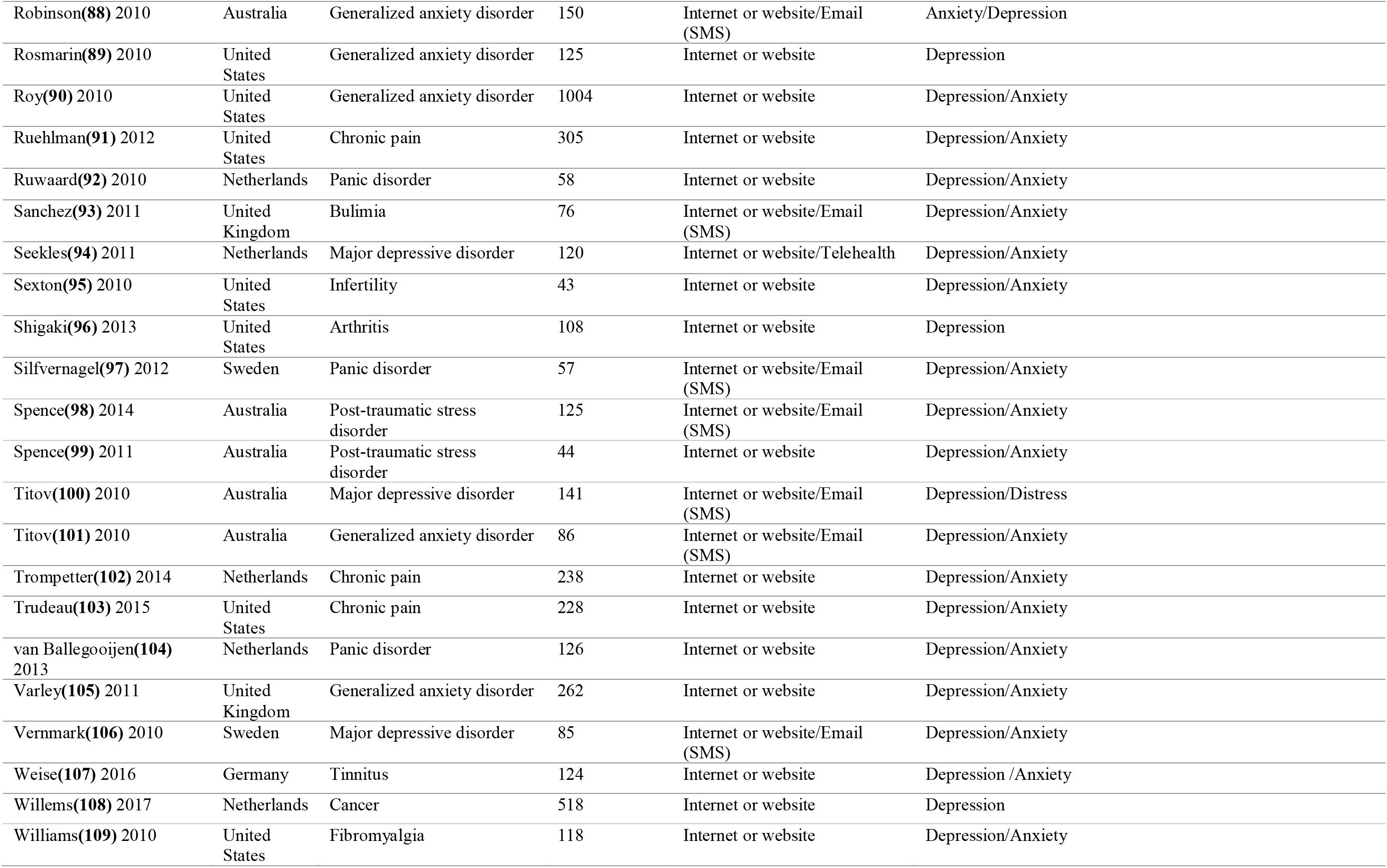
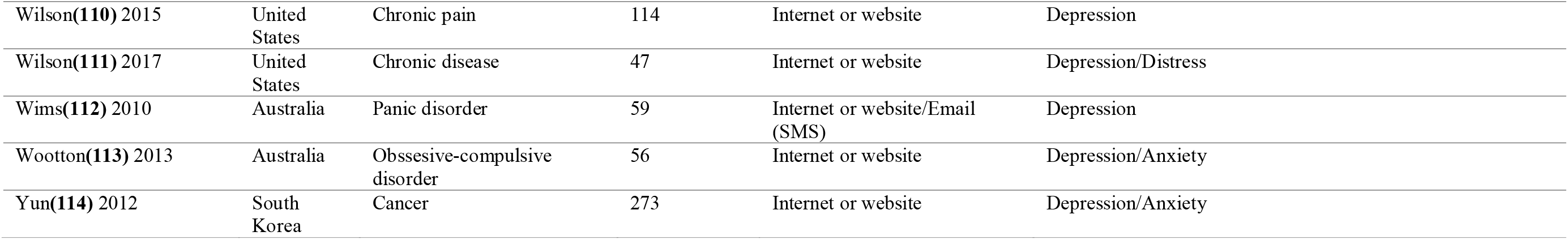
DESCRIPTION OF INCLUDED STUDIES IN THE META-ANALYSIS.

A summary of the estimated effect size for each intervention characteristics, heterogeneity and inconsistency for all comparisons, and for both outcomes are presented in Table 2.

**Table 2.** Summary of comparative intervention caracteristics

| Interventions characteristics | n participants | Effect estimate (95% CI) | | Heterogeneity, $\chi^2$<br>(p value) | Inconsistency,<br>$I^2$ (%) |
| --- | --- | --- | --- | --- | --- |
| *Anxiety outcomes* |  |  |  |  |  |
| Mobile Applications | 76 | -0,52 | (-0,04, -1,00) | N/A | N/A |
| Digital Video Disc (other) | 220 | -0,15 | (0,11, -0,42) | N/A | N/A |
| Computer Software | 114 | -0,57 | (-0,18, -0,96) | 3.08 (0.08) | 67.55 |
| Connected Devices | 51 | -0,22 | (0,33, -0,77) | N/A | N/A |
| Electronic Messaging | 2700 | -0,48 | (-0,39, -0,56) | 95.30 (< .0005) | 79.01 |
| Internet / Website | 8305 | -0,39 | (-0,35, -0,44) | 396.02 (< .0005) | 85.61 |
| Telehealth / Telemedecine | 394 | -0,50 | (-0,29, -0,70) | 21.48 (< .0005) | 86.03 |
| Virtual Reality | 80 | -1,73 | (-1,21, -2,25) | N/A | N/A |
| Self-administred | 5312 | -0,35 | (-0,30, -0,41) | 231.19 (< .0005) | 87.46 |
| Partially guided | 3206 | -0,46 | (-0,39, -0,53) | 155.93 (< .0005) | 82.04 |
| Guided | 201 | -0,46 | (-0,16, -0,76) | 36.55 (< .0005) | 94.53 |
| Overall effect | 8719 | -0,40 | (-0,35, -0,44) | 428.75 (< .0005) | 85.77 |
| *Depression outcomes* |  |  |  |  |  |
| Mobile Applications | 196 | -0,26 | (0,02, -0,55) | 0.09 (0.768) | 0 |
| Digital Video Disc (other) | 273 | -0,10 | (0,14, -0,34) | 1.41 (0.241) | 29.01 |
| Computer Software | 191 | -0,55 | (-0,26, -0,85) | 5.71 (0.134) | 47.42 |
| Connected Devices | 51 | -0,13 | (0,42, -0,68) | N/A | N/A |
| Electronic Messaging | 2915 | -0,48 | (-0,40, -0,56) | 146.62 (< .0005) | 83.63 |
| Internet / Website | 9492 | -0,33 | (-0,29, -0,37) | 299.91 (< .0005) | 76.99 |
| Telehealth / Telemedecine | 394 | -0,75 | (-0,54, -0,96) | 15.18 (0.002) | 80.24 |
| Self-administred | 6379 | -0,28 | (-0,23, -0,33) | 113.53 (< .0005) | 65.65 |
| Partially Guided | 3470 | -0,43 | (-0,36, -0,50) | 176.59 (< .0005) | 81.88 |
| Guided | 121 | 0,16 | (0,53, -0,22) | 4.96 (0.026) | 79.82 |
| Depression overall effect | 9970 | -0,33 | (-0,29, -0,37) | 312.90 (< .0005) | 76.35 |

CI: Confidence interval

### Global mean differences

The first analysis aimed at the global group differences of any digital intervention compared to usual care or another digital intervention to manage anxiety or depression for people with any concomitant chronic condition.

A total of 62 studies including 8719 participants with anxiety outcomes measures were included in the meta-analysis of the overall effect of digital interventions on anxiety outcomes (additional file 1). The results showed a significant decrease in the anxiety score related to digital health interventions compared to usual care or another digital intervention [Standardized Mean Difference (SMD) = -0.40; 95% Confidence Interval (CI) = -0.35; -0.44)]. Although heterogeneity is high between studies (I^2^ = 85.77%) the results are relatively consistent across studies (Table 2).

Regarding depression outcomes, we conducted a meta-analysis including 75 studies with 9970 participants. Figure 4 shows a significant reduction in depression scores associated with the use of digital health interventions compared to usual care (SDM = -0.33; 95% CI [-0.29,-0.37]). Heterogeneity is also high for this overall comparison (I^2^ = 76.35%) with results consistent across studies (Table 2).

**Figure 4.**
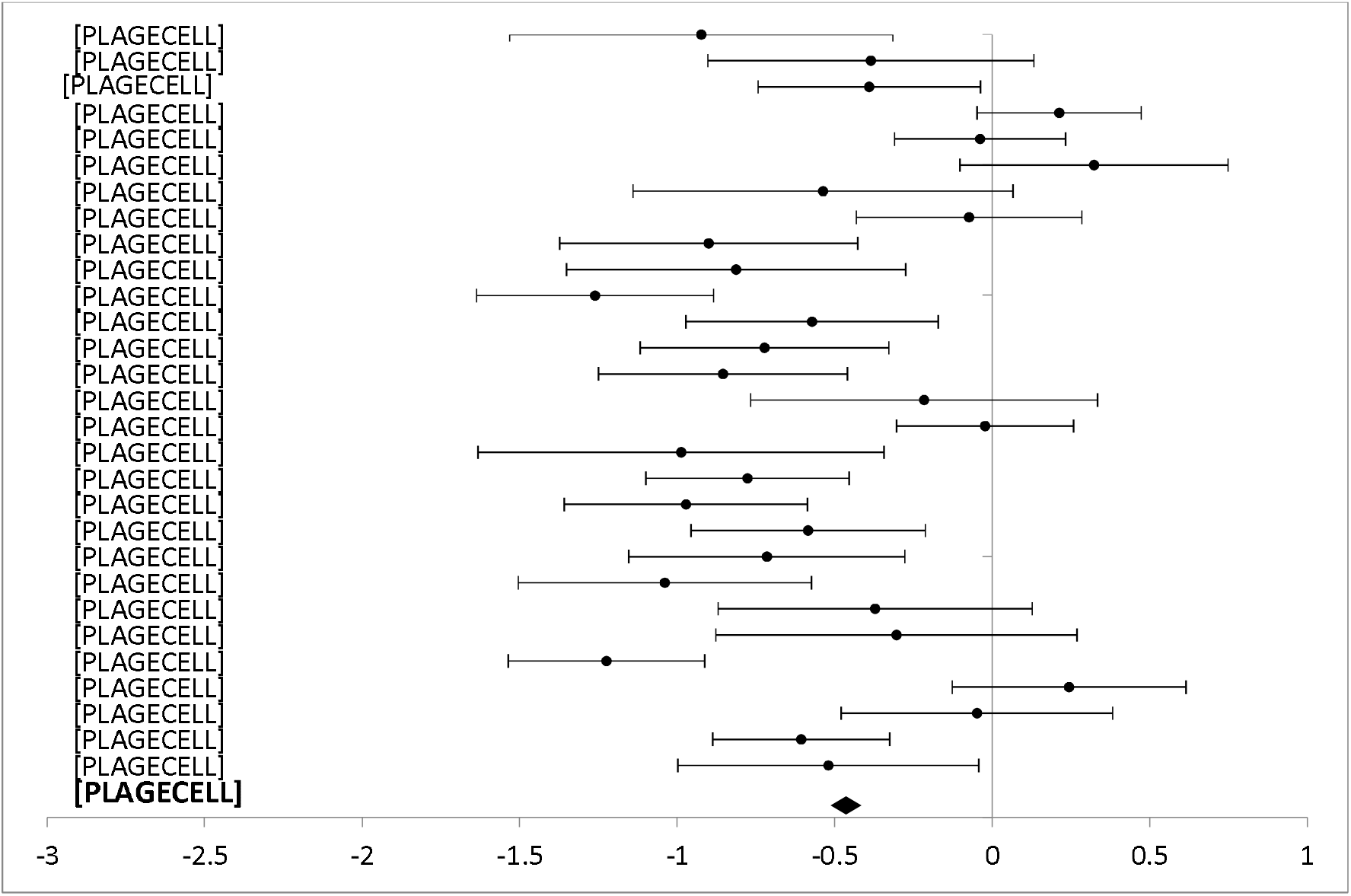
Forest plot: Subgroup analysis of partially guided interventions for any digital intervention vs. usual care or another digital intervention to manage anxiety in people with any concomitant chronic condition.

#### Level of professional support

The forest plots for comparing the level of professional support are presented for anxiety (figures 3 and 4) and depression (figures 5 et 6) outcomes separately.

**Figure 5.**
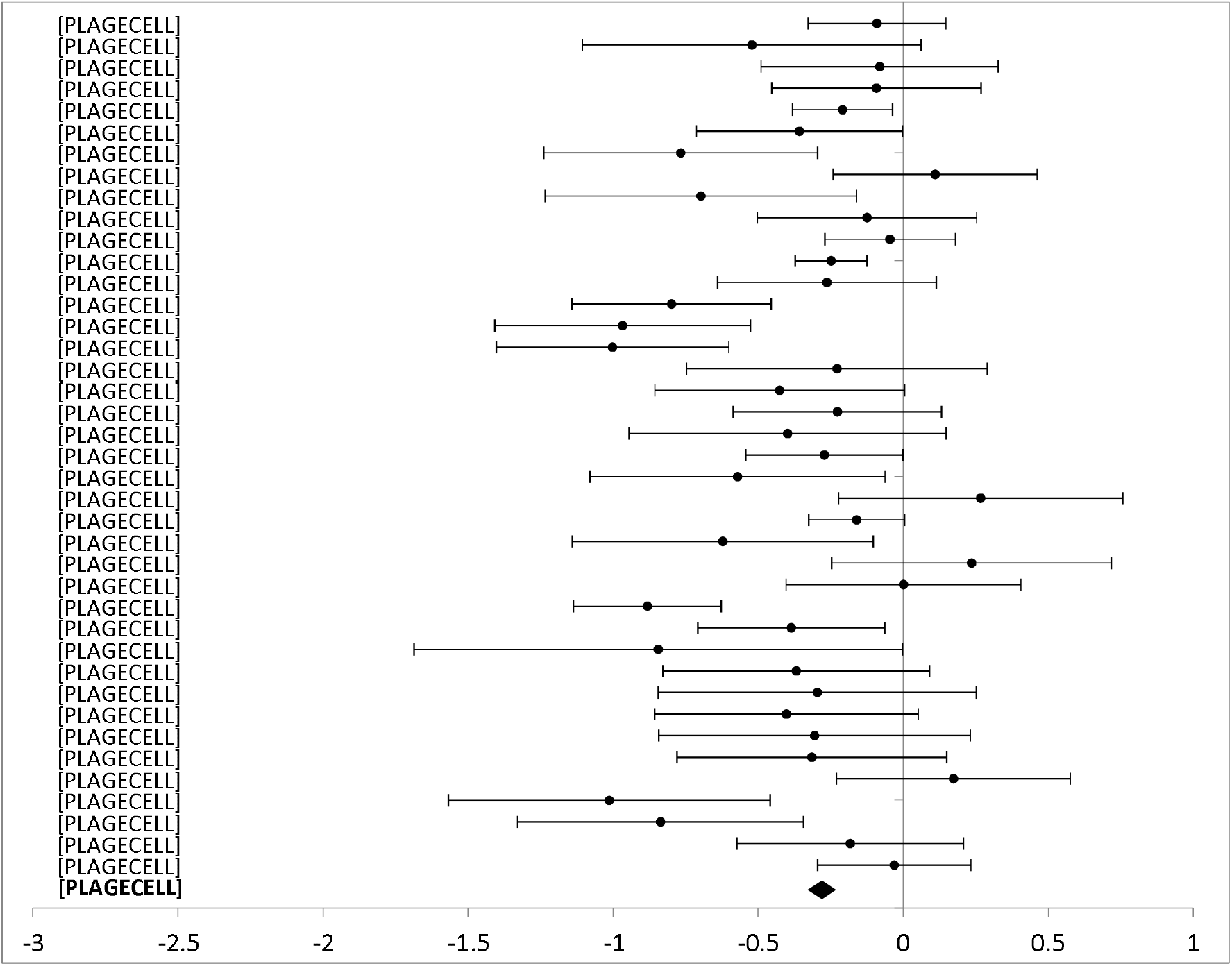
Forest plot: Subgroup analysis of self-directed interventions for any digital intervention vs. usual care or another digital intervention to manage depression in people with any concomitant chronic condition.

**Figure 6.**
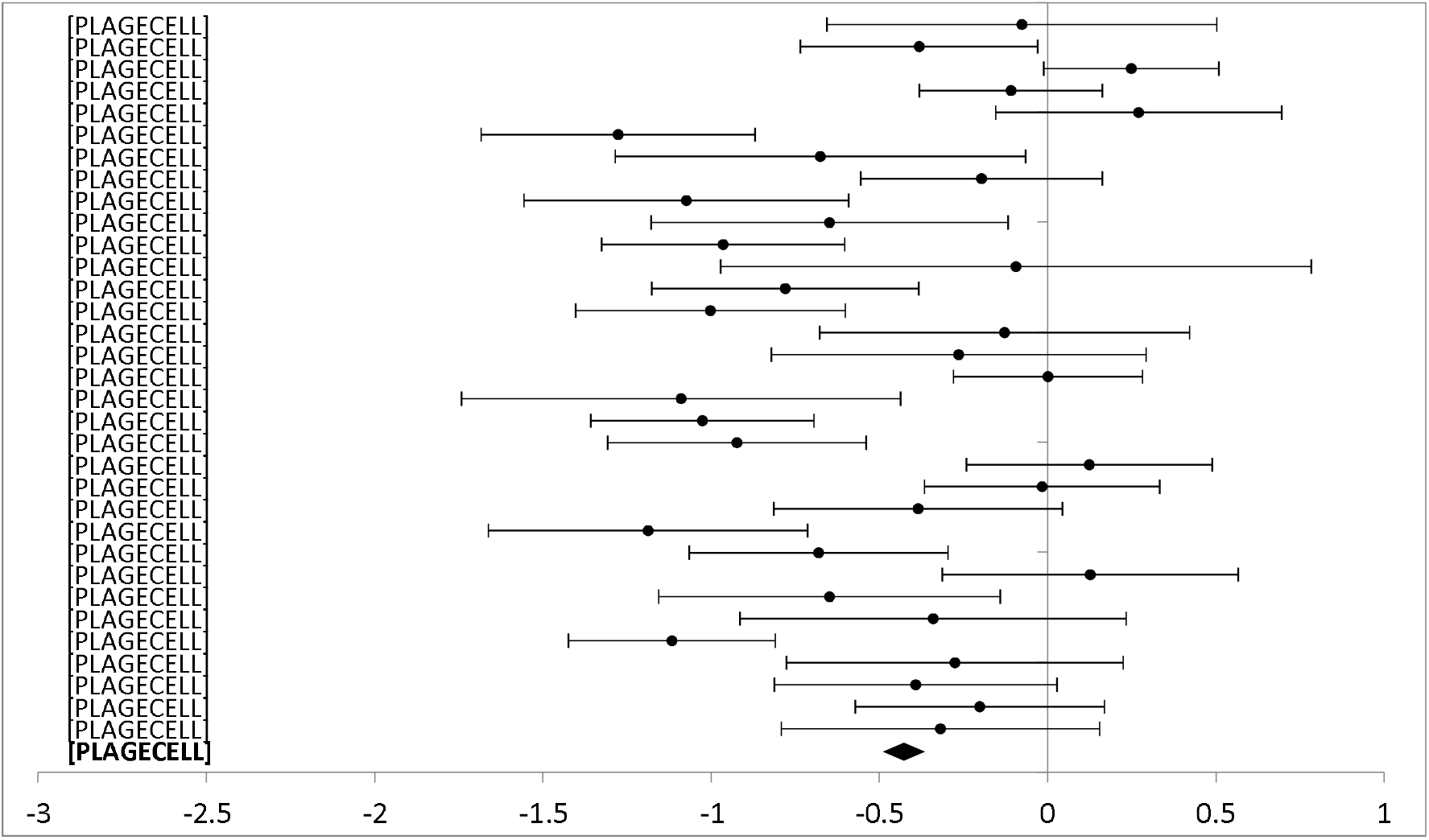
Forest plot: Subgroup analysis of partially guided interventions for any digital intervention vs. usual care or another digital intervention to manage depression in people with any concomitant chronic condition.

Completely self-administered was the delivery method used in 30 studies (5312 participants) with anxiety outcomes and 40 studies (6379 participants) reporting depression outcomes. Self-administered delivery showed a significant decrease in anxiety scores [SMD -0.35 (−0.30, -0.41)] and depression scores [(SMD -0.28 (−0.23, -0.33)].

Partial support by a healthcare provider was used in 29 studies (3206 participants) reporting anxiety outcomes, and 33 studies (3470 participants) reporting depression outcomes. Partial support showed a significant decrease in anxiety [SMD -0,46 (−0.39, -0.53)] and depression scores [SMD -0,43 (−0.36, - 0.50)].

Interventions entirely guided by a healthcare professional were used in three studies (201 participants) reporting anxiety outcomes and two studies (121 participants) reporting depression outcomes. Interventions entirely supported by healthcare professionals showed a significant difference for anxiety scores [SMD -0.46 (−0.16, -0.76)] but no significant difference between groups for depression scores [SMD 0.15 (0.53, -0.21)].

#### Type of technology

Eight different technologies were used in studies reporting anxiety outcomes. and half had more than one study on which we could perform a meta-analysis (figure 7 and 8). For studies reporting a depression outcome, seven technologies were used, and five of them had more than one study (figure 9 and 10). Forest plots that include all studies are presented in additional file 3 and 4.

**Figure 7.**
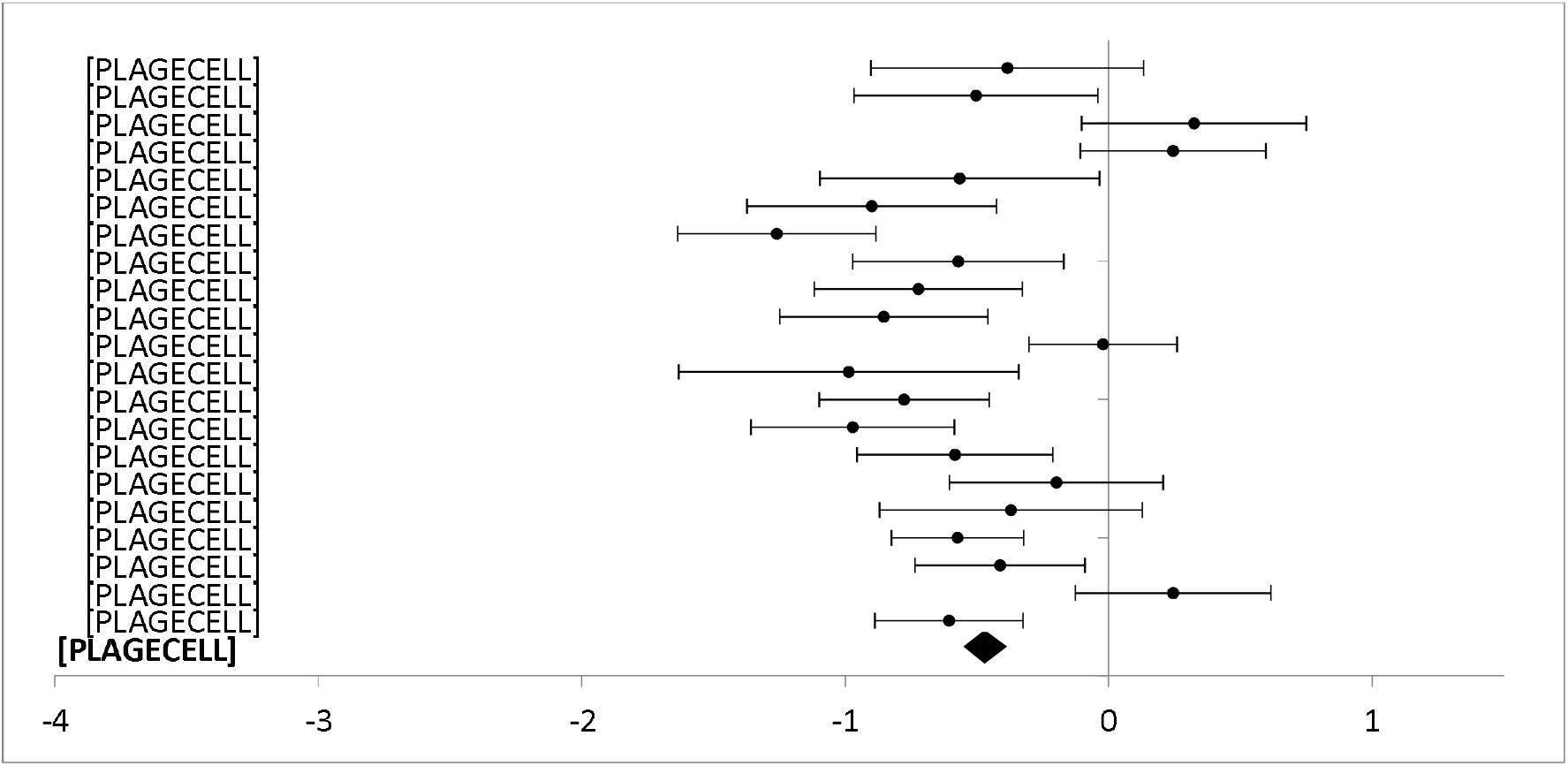
Forest plot: Subgroup analysis of electronic messaging interventions for any digital intervention vs. usual care or another digital intervention to manage anxiety in people with any concomitant chronic condition.

**Figure 8.**
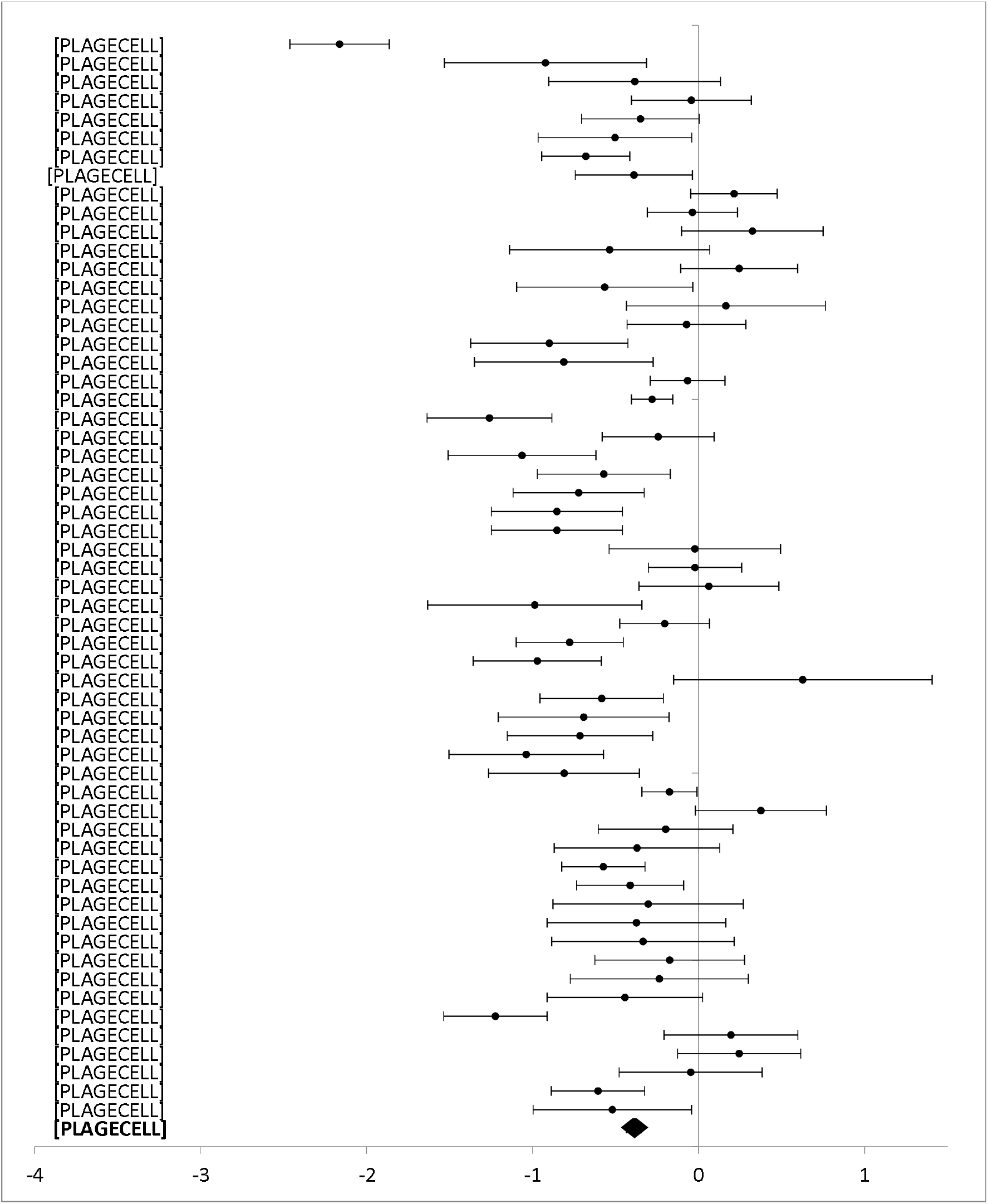
Forest plot: Subgroup analysis of internet or website interventions for any digital intervention vs. usual care or another digital intervention to manage anxiety in people with any concomitant chronic condition.

**Figure 9.**
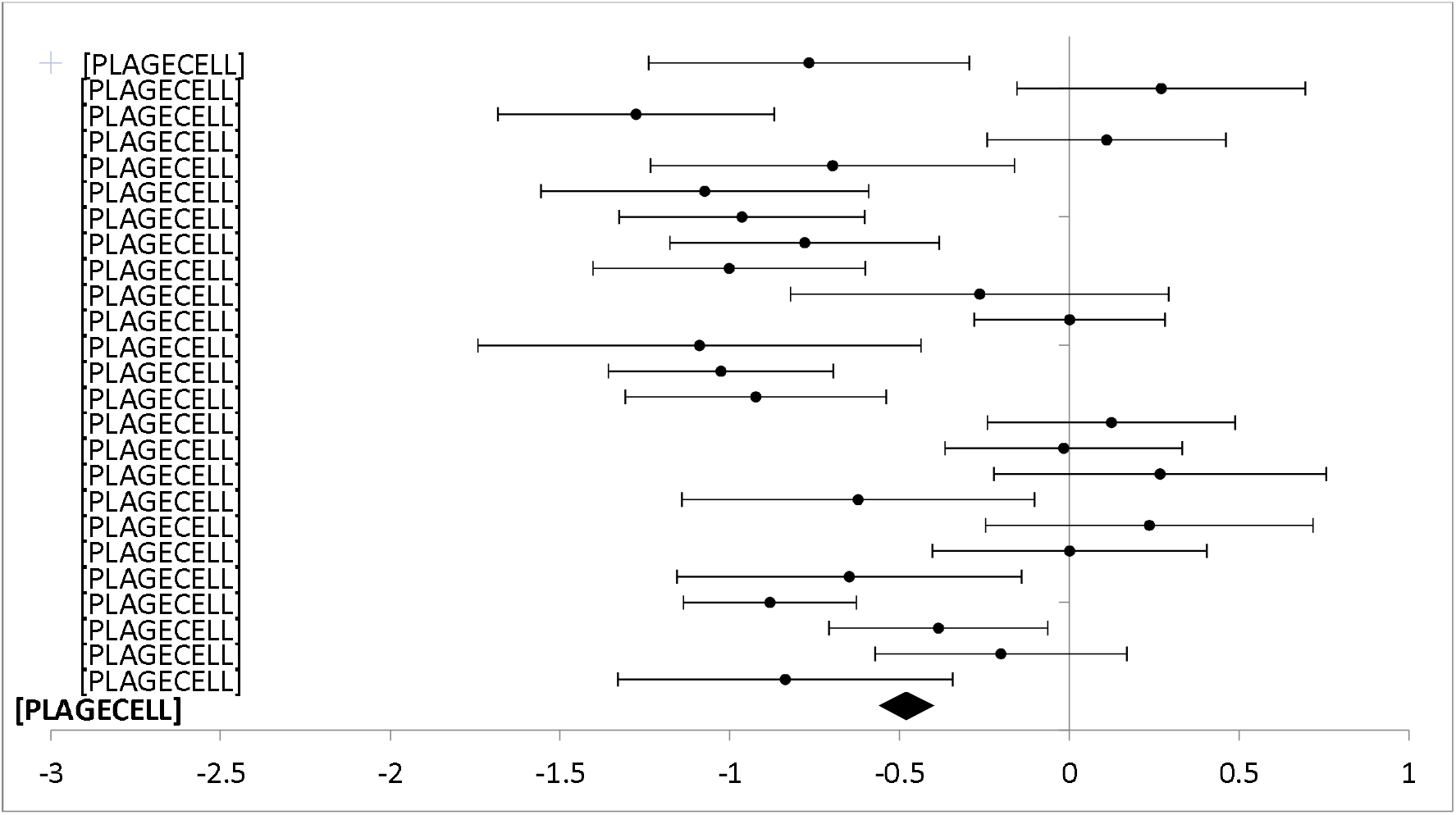
Forest plot: Subgroup analysis on electronic messaging interventions for any digital intervention vs. usual care or another digital intervention to manage depression in people with any concomitant chronic condition.

**Figure 10.**
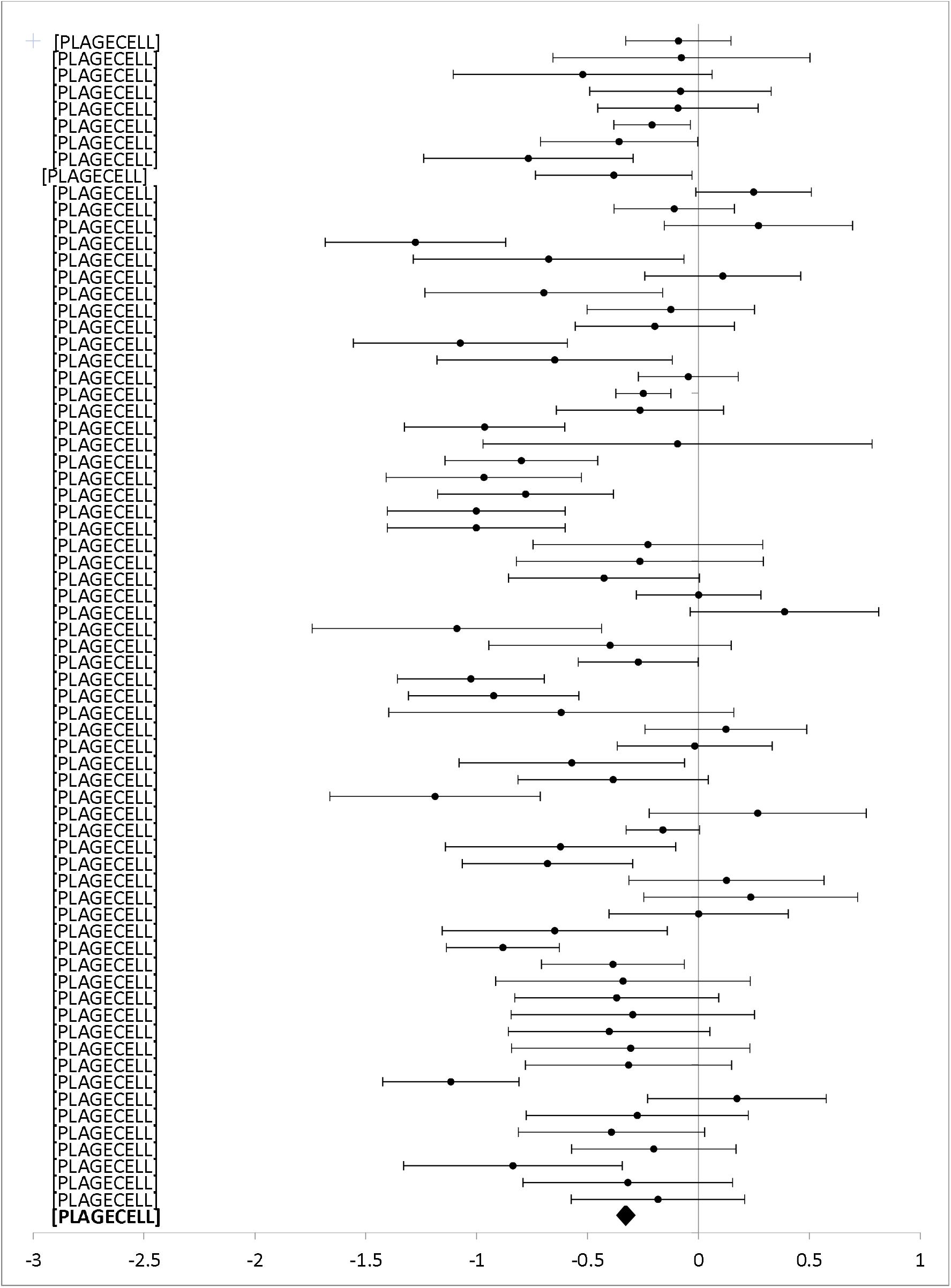
Forest plot: Subgroup analysis on internet and website interventions for any digital intervention vs. usual care or another digital intervention to manage depression in people with any concomitant chronic condition.

Electronic messaging was used in 23 studies (2700 participants) reporting anxiety outcomes, and 25 studies (2915 participants) reporting depression outcomes. Electronic messaging showed significant improvement in anxiety scores [SMD -0.48 (−0.39, -0.56)] and depression scores [SMD -0.48 (−0.40, - 0.56)].

Internet and website technologies were used in 58 studies (8305 participants) reporting anxiety outcomes, and 70 studies (9492 participants) reporting depression outcomes. Internet and website interventions showed significant improvement in anxiety [SMD -0.39 (−0.35, -0.44)] and depression scores [SMD -0.33 (−0.29, -0.37)].

Telehealth and telemedicine were used in four studies (394 participants) reporting anxiety and/or depression outcomes. Telehealth and telemedicine showed significant improvement in anxiety [SMD -0.50 (−0.29, -0.70)] and depression scores [SMD -0.75 (−0.54, -0.96)].

A computer software was used in two studies (114 participants) reporting anxiety outcomes and four studies (191 participants) with depression outcomes. Computer software showed significant improvement in anxiety [SMD -0.57 (−0.18, -0.96)] and depression scores [SMD -0.55 (−0.26, -0.85)].

Mobile applications were used in one study (76 participants) with anxiety outcomes and two studies (196 participants) with depression outcomes. Mobile applications showed significant improvement for anxiety scores [SMD -0.52 (−0.04, -1.00)], but no significant difference between groups for depression scores [SMD-0.26 (0.02, -0.55)].

Connected devices were used in one study with 51 participants and showed no differences between groups for anxiety [SMD -0.22 (0.33, -0.77)] and depression scores [SMD -0.13 (0.42, -0.68)]

Virtual reality was used in one study with 80 participants with anxiety outcomes and showed significant improvement on anxiety scores [SMD -1.73 (−1.21, -2.25)].

One other type of technology in the form of Digital Video Disk (DVD) was used in one study (220 participants) with anxiety outcomes and two studies (273 participants) with depression outcomes. DVD interventions showed no differences between groups for anxiety [SMD -0.15 (0.11, -0.42)] and depression scores [SMD -0.10 (0.14, -0.34)].

### Risk of Bias

Figure 11 presents the risk of bias across studies for each domain. Most of the included studies showed an overall low risk of bias. However, risk of bias was generally high for the Domain 4: *Risk of bias in the measurement of the outcomes*. In fact, blinding of study participants was not done in most studies and outcomes were self-reported, leading to a high risk of performance bias. This bias is present across studies and would eventually lead to an overestimation of the effect.

**Figure 11.**
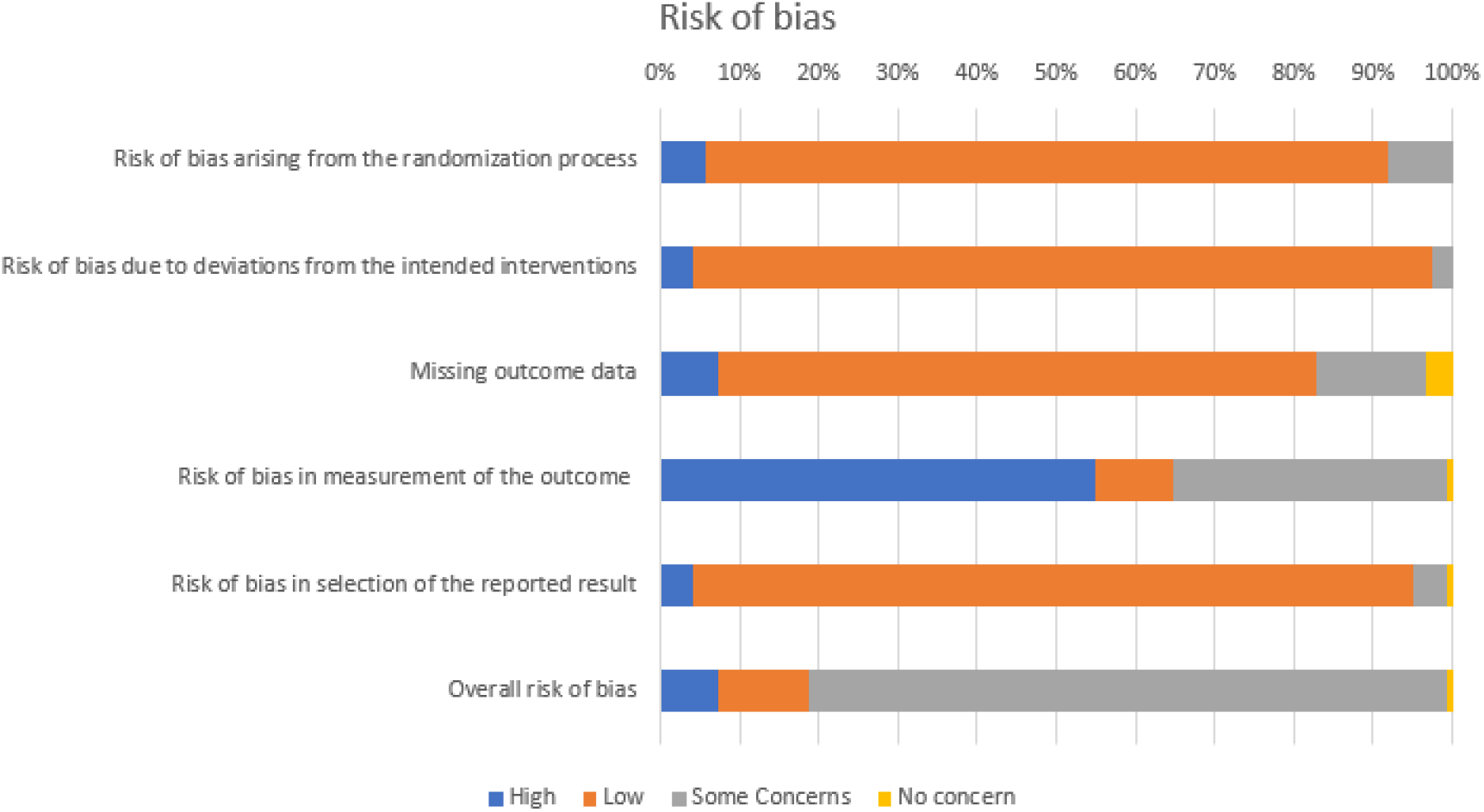
Assessment of the risk of bias across studies

## DISCUSSION

This knowledge synthesis aimed to rapidly provide evidence for knowledge users regarding the types of digital mental health interventions that were the most effective for people living with a concomitant chronic disease. This secondary analysis answers a specific research question based on knowledge users’ needs and prioritization. Thus, preliminary work in the form of a workshop and a two-round Delphi study were conducted to identify the top-priority question for knowledge users. This question was: “*What types of digital health interventions are the most effective for the management of concomitant mental health and chronic disease conditions in adults*?”. A total of 84 primary studies including anxiety and depression outcomes were identified from the systematic reviews.

Overall, the results show that digital health interventions are effective to manage mental health issues in adults living with a concomitant chronic condition. The magnitude of the effect varies for anxiety and depression, and heterogeneity is generally high, but the effect size and direction are consistent across studies. Subgroup analyses show that digital mental health interventions with partial support from a healthcare provider have a larger effect size than self-administered interventions. These results are in line with what is known for other populations (21, 22). There is not enough evidence to conclude in the effectiveness of digital interventions that are completely guided by a healthcare professional because of the lack of studies in this category. One key challenge of self-administered digital mental health interventions is sustaining engagement and reducing dropout (23, 24). Partially supported interventions are mitigating these challenges by improving interactivity and personalization (25, 26). However, it is also documented that patients could prefer to only interact with a platform instead than talking with a healthcare professional, highlighting a need for flexible interventions (27).

Regarding the type of technology used, our analyses show that the most effective type of intervention is electronic messaging, but that all types of technologies are effective for both anxiety and depression scores. This finding adds to the literature on the use of digital mental health to reduce disparities. Indeed, as our results show that all types of technologies are equally or more effective than usual care, stakeholders could choose and implement interventions in relation with the needs of the population. For example, decision makers can tailor their choices with respect to cost, ease of access or easing stigma barriers (28).

More research will be needed for newer technologies, such as mobile apps and virtual reality, which have showed effectiveness only in a small number of studies with a large confidence interval.

The significant heterogeneity observed between studies in every comparison is not surprising and could be likely due to differences in comorbidity, outcome measure used, and content of the intervention. However, patterns shown in this meta-analysis are useful for clinical use and implementation.

This knowledge synthesis was informed by knowledge users in order to validate the review questions considering their needs, and identify knowledge gaps that would require more evidence. We used a two-stage process, starting with a rapid review of systematic reviews followed by a secondary analysis of the primary studies. Although we used a systematic approach for selecting these studies, a major limitation is that more recent studies were not included in the analyses. In order to meet the requirement of the funding agency and the urgent need for evidence in the current pandemic, we considered only the most recent studies (published from 2010) from the included reviews. We also assessed the risk of bias in the included studies.

Results from the meta-analyses should be interpreted with caution since heterogeneity was generally high. Further analyses, including subgroup analyses for different populations are needed in order to provide a more detailed and nuanced portrait of the effectiveness of digital mental health interventions. Furthermore, sensitivity analyses, notably by considering the risk of bias related to the lack of blinding of participants, would be required to minimize the risk of an overestimation of the effect. Finally, we cannot rule out the possibility of publication bias, as well as other factors that could lessen the level of confidence in the reported effects.

Available evidence suggests that digital health interventions such as internet-based cognitive behavioural therapy (iCBT) could be effective and provide an alternative to face-to-face psychological interventions to manage mental health issues in adults living with a concomitant chronic condition. In the context of the COVID-19 pandemic, digital technologies have played a key role in healthcare. Many of these innovations support the care of people in need of medical attention, including those with chronic illnesses. In response to the current crisis, but also to better prepare for the post-crisis and future crises, digital mental health interventions could be a useful tool to manage mental health problems in people living with chronic conditions.

## CONCLUSION

This knowledge synthesis provides an overview of the current evidence regarding the use of digital health interventions to improve mental health in people living with a chronic condition. Knowledge users’ most urgent need was for evidence on which type of digital interventions to use for mental health management. While our meta-analysis indicates different levels of effectiveness associated with digital interventions’ characteristics, all technologies and levels of support can be used with consideration of implementation context and population.

## Data Availability

The datasets used and/or analyzed during the current study are available from the corresponding author on reasonable request.

## DECLARATIONS

### Ethics approval and consent to participate

Not applicable

### Consent for publication

Not applicable

## Authors’ contributions

Maxime Sasseville, Annie LeBlanc, Marie-Pierre Gagnon, Maud-Christine Chouinard, Marianne Beaulieu, Nicolas Beaudet, Pascale Cholette Christine Aspiros, Alain Larouche, Guylaine Chabot identified the need for this study and contributed to its conception and design. Maxime Sasseville, Marie-Pierre Gagnon, Mylène Boucher, Michèle Dugas, Mbemba Gisèle, Jack Tchuente and Romina Barony conducted the data collection. Jack Tchuente and Maxime Sasseville performed the data analysis. Maxime Sasseville developed the first draft of the report. All authors contributed to writing and editing and gave the final approval of the version submitted.

## Acknowledgements

The authors would like to thank the knowledge users for their support throughout this review and the SPOR Evidence Alliance for their support.

---

**Figure 1.**
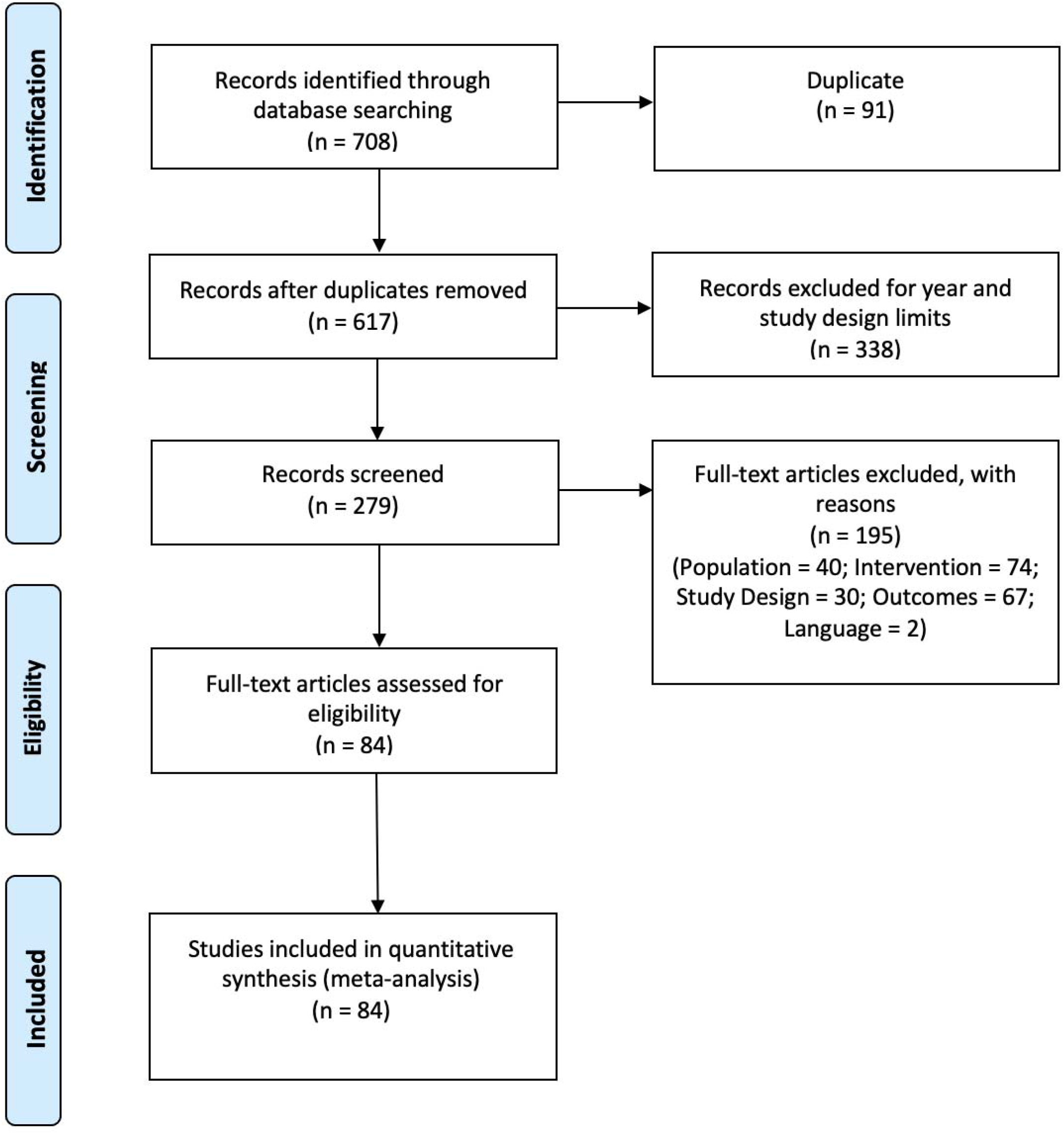
Study Flow Diagram: Meta-analysis Moher D, Liberati A, Tetzlaff J, Altman DG, The PRISMA Group (2009). Preferred Reporting Items for Systematic Reviews and Meta-Analyses: The PRISMA Statement. PLoS Med 6(6): e1000097. doi:10.1371/journal.pmed1000097

Additional file 1 : Forest plot: Subgroup analysis of self-directed interventions for any digital intervention vs. usual care or another digital intervention to manage anxiety in people with any concomitant chronic condition.

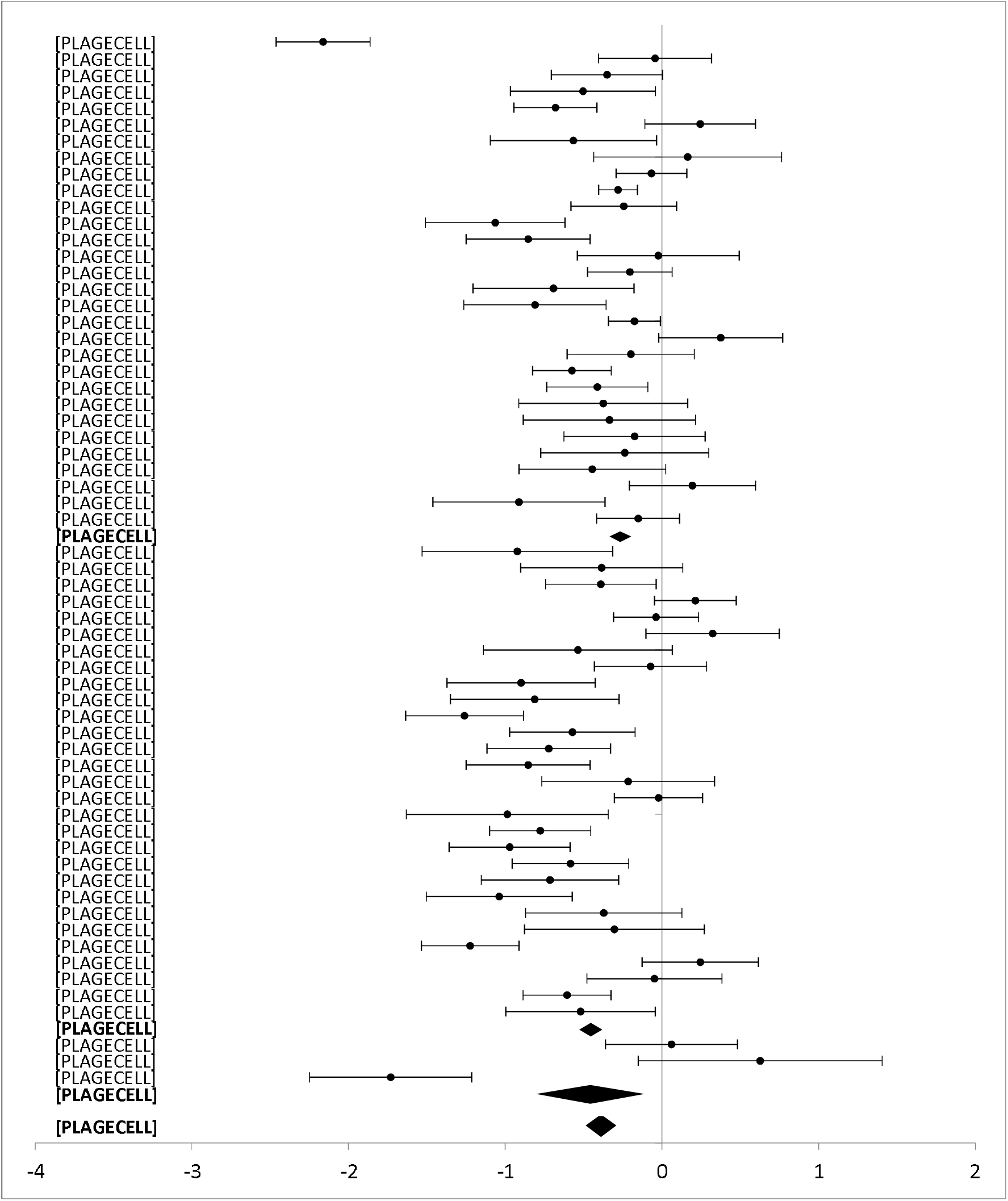

*Additional file 2: Forest plot: Subgroup analysis on level of professional support for any digital intervention vs. usual care or another digital intervention to manage depression in people with any concomitant chronic condition*.

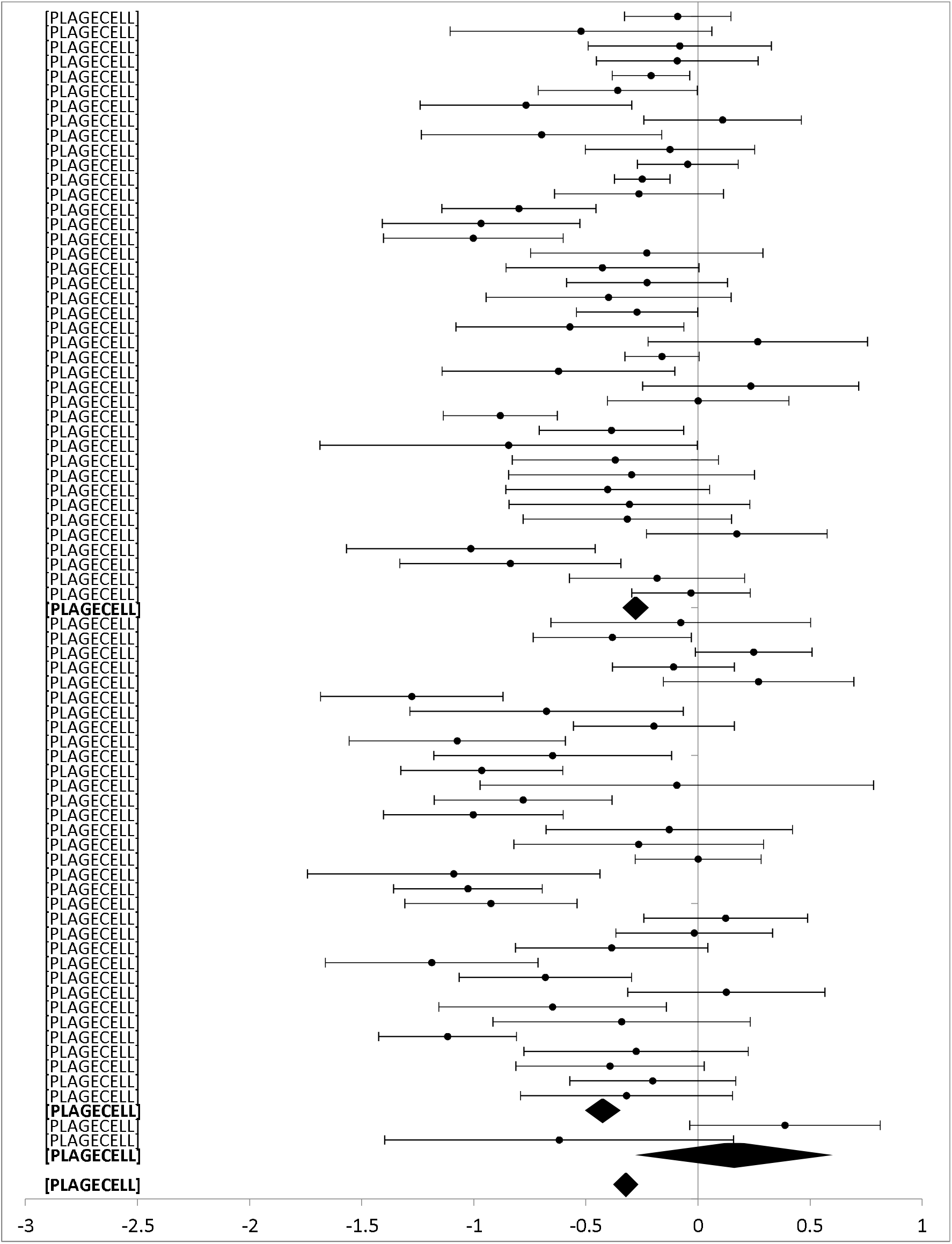

Additional file 3. *Forest plot: Subgroup analysis on type of technology for any digital intervention vs. usual care or another digital intervention to manage anxiety in people with any concomitant chronic condition*.

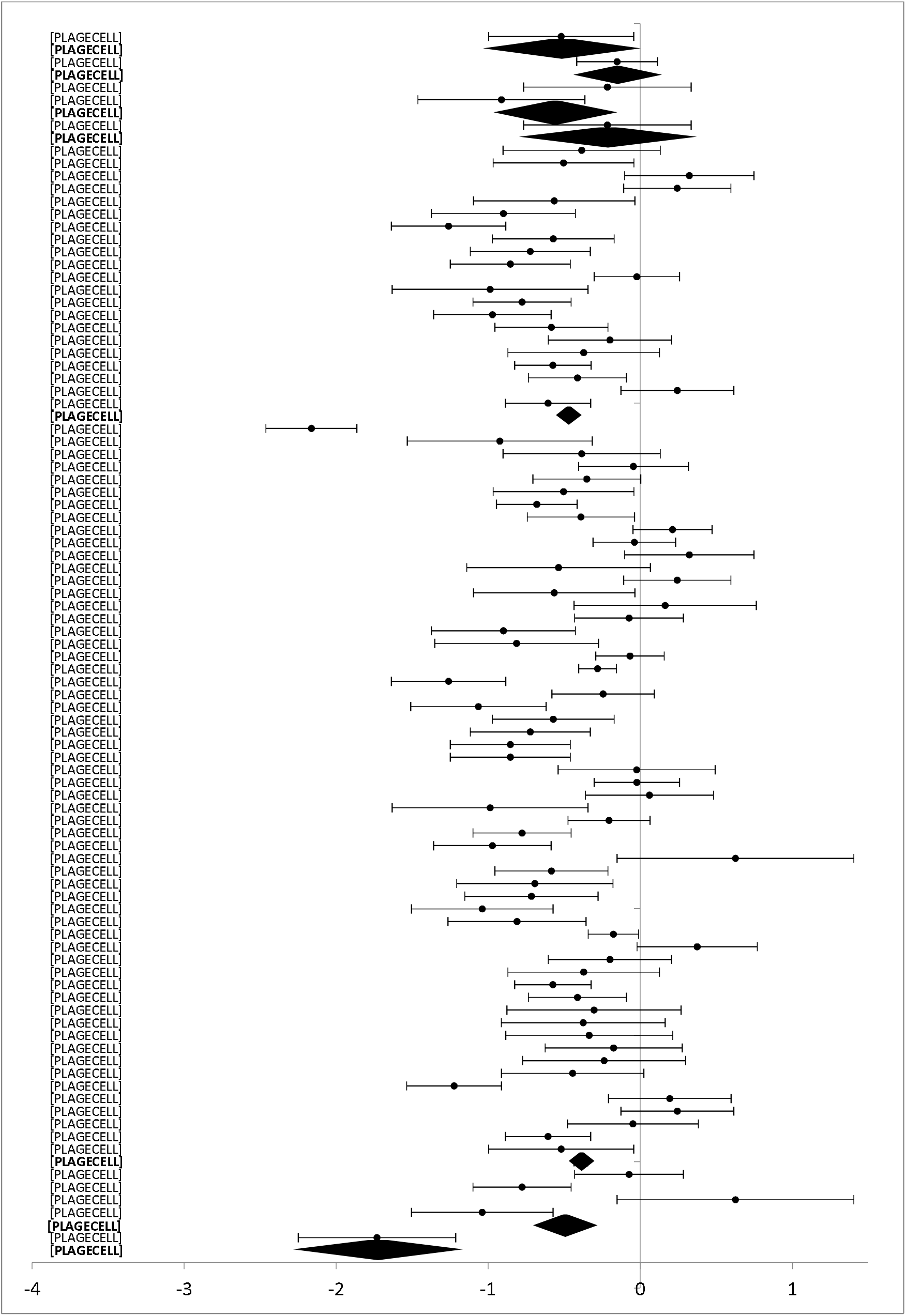

Additional file 4. *Forest plot: Subgroup analysis on type of technology for any digital intervention vs. usual care or another digital intervention to manage depression in people with any concomitant chronic condition*.

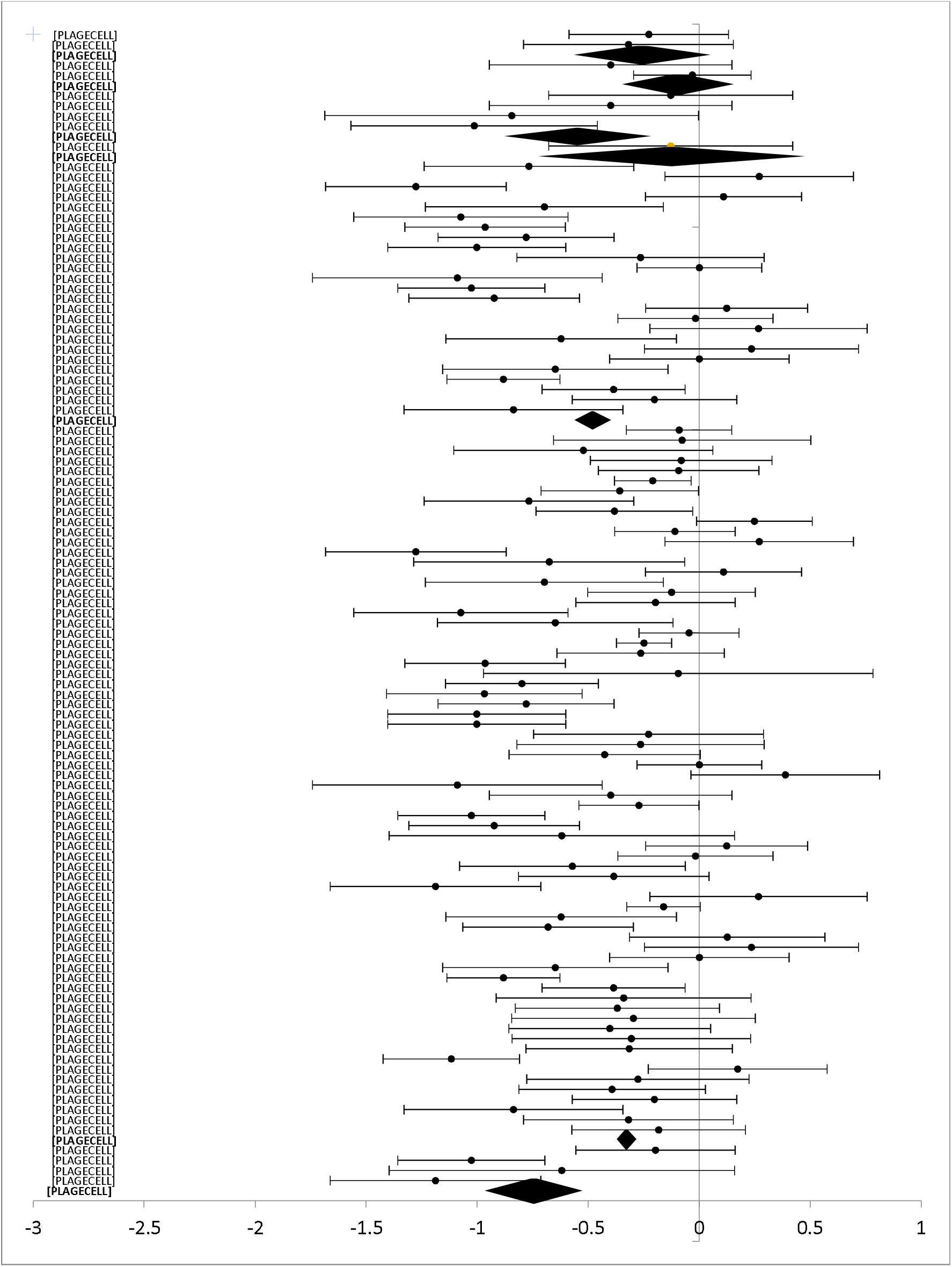

## REFERENCES

1. World Health Organisation. Global status report on noncommunicable diseases. Geneva, Switzerland; 2014.

2. DeJean D, Giacomini M, Vanstone M, Brundisini F. Patient experiences of depression and anxiety with chronic disease: a systematic review and qualitative meta-synthesis. Ont Health Technol Assess Ser. 2013;13(16):1–33.

3. Watson LC, Amick HR, Gaynes BN, Brownley KA, Thaker S, Viswanathan M, et al. Practice-based interventions addressing concomitant depression and chronic medical conditions in the primary care setting: a systematic review and meta-analysis. J Prim Care Community Health. 2013;4(4):294–306.

4. Sherbourne CD, Wells KB, Meredith LS, Jackson CA, Camp P. Comorbid anxiety disorder and the functioning and well-being of chronically ill patients of general medical providers. Arch Gen Psychiatry. 1996;53(10):889–95.

5. Stein MB, Cox BJ, Afifi TO, Belik SL, Sareen J. Does co-morbid depressive illness magnify the impact of chronic physical illness? A population-based perspective. Psychol Med. 2006;36(5):587-96.

6. Donnelly C, Ashcroft R, Bobbette N, Mills C, Mofina A, Tran T, et al. Interprofessional Primary Care During COVID-19: The Provider Perspective. Available at SSRN 3629453. 2020.

7. García-Lizana F,Muñoz-Mayorga I. Telemedicine for depression: a systematic review. Perspect Psychiatr Care. 2010;46(2):119–26.

8. Richards D, Richardson T. Computer-based psychological treatments for depression: a systematic review and meta-analysis. Clin Psychol Rev. 2012;32(4):329–42.

9. Lattie EG, Adkins EC, Winquist N, Stiles-Shields C, Wafford QE, Graham AK. Digital mental health interventions for depression, anxiety, and enhancement of psychological well-being among college students: systematic review. Journal of medical Internet research. 2019;21(7):e12869.

10. Garritty C, Gartlehner G, Nussbaumer-Streit B, King VJ, Hamel C, Kamel C, et al. Cochrane Rapid Reviews Methods Group offers evidence-informed guidance to conduct rapid reviews. Journal of Clinical Epidemiology. 2020.

11. Sasseville M, LeBlanc A, Boucher M, Dugas M, Mbemba G, Tchuente J, et al. Digital health interventions for the management of mental health in people with chronic diseases: a rapid review.. BMJ Open. 2021.

12. Garritty C GG, Kamel C, King VJ, Nussbaumer-Streit B, Stevens A, Hamel C, Affengruber L. Cochrane Rapid Reviews. Interim Guidance from the Cochrane Rapid Reviews Methods Group. March 2020.

13. Higgins J TJ, Chandler J, Cumpston M, Li T, Page M, et al. Cochrane Handbook for Systematic Reviews of Interventions Version 6.0 (Updated July 2019). 2019.

14. Gagnon M-P, Sasseville M, LeBlanc A. A stepped approach for knowledge synthesis to rapidly inform clinical decisions: methods and lessons learned. Science of Nursing and Health Practices. Submitted

15. Harris PA, Taylor R, Minor BL, Elliott V, Fernandez M, O’Neal L, et al. The REDCap consortium: Building an international community of software platform partners. Journal of biomedical informatics. 2019;95:103208.

16. Evidence Partners. DistillerSR [Computer Program]. Ottawa, Canada: Evidence Partners. 2011.

17. World Health O. Classification of digital health interventions v1.0: a shared language to describe the uses of digital technology for health. Geneva: World Health Organization; 2018 2018. Contract No.: WHO/RHR/18.06.

18. Liverpool S, Mota CP, Sales CMD, Čuš A, Carletto S, Hancheva C, et al. Engaging Children and Young People in Digital Mental Health Interventions: Systematic Review of Modes of Delivery, Facilitators, and Barriers. J Med Internet Res. 2020;22(6):e16317.

19. Hollis C, Falconer CJ, Martin JL, Whittington C, Stockton S, Glazebrook C, et al. Annual Research Review: Digital health interventions for children and young people with mental health problems - a systematic and meta-review. J Child Psychol Psychiatry. 2017;58(4):474–503.

20. Moher D, Liberati A, Tetzlaff J, Altman DG. Preferred reporting items for systematic reviews and meta-analyses: the PRISMA statement. PLoS Med. 2009;6(7):e1000097.

21. Schueller SM, Tomasino KN, Mohr DC. Integrating human support into behavioral intervention technologies: the efficiency model of support. Clinical Psychology: Science and Practice. 2017;24(1):27–45.

22. Wright JH, Owen JJ, Richards D, Eells TD, Richardson T, Brown GK, et al. Computer-assisted cognitive-behavior therapy for depression: a systematic review and meta-analysis. The Journal of clinical psychiatry. 2019;80(2):0-.

23. Eysenbach G. The law of attrition. Journal of medical Internet research. 2005;7(1):e11.

24. Karapanos E. Sustaining user engagement with behavior-change tools. Interactions. 2015;22(4):48–52.

25. Doherty G, Coyle D, Sharry J, editors. Engagement with online mental health interventions: an exploratory clinical study of a treatment for depression. Proceedings of the SIGCHI Conference on Human Factors in Computing Systems; 2012.

26. Rennick-Egglestone S, Knowles S, Toms G, Bee P, Lovell K, Bower P, editors. Health Technologies’ In the Wild’ Experiences of Engagement with Computerised CBT. Proceedings of the 2016 CHI Conference on Human Factors in Computing Systems; 2016.

27. Bradford S, Rickwood D. Young people’s views on electronic mental health assessment: prefer to type than talk? Journal of child and family studies. 2015;24(5):1213–21.

28. Ralston AL, Andrews III AR, Hope DA. Fulfilling the promise of mental health technology to reduce public health disparities: Review and research agenda. Clinical Psychology: Science and Practice. 2019;26(1):e12277.

29. Moher D, Liberati A, Tetzlaff J, Altman DG, Group P. Preferred reporting items for systematic reviews and meta-analyses: the PRISMA statement. J Clin Epidemiol. 2009;62(10):1006–12.

30. Aguado L, Claudia X, Taylor Teletia R, McMillan, Susan, et al. Use and Helpfulness of Self-Administered Stress Management Therapy in Patients Undergoing Cancer Chemotherapy in Community Clinical Settings. Journal of Psychosocial Oncology. 2012;30(1):57–80.

31. Andersson, E, Enander, J, Andren, P, et al. Internet-based cognitive behaviour therapy for obsessive-compulsive disorder: a randomized controlled trial. Psychol Med. 2012;42(10):2193-203.

32. Andersson, Gerhard, Carlbring, Per, Furmark, Tomas, et al. Therapist Experience and Knowledge Acquisition in Internet-Delivered CBT for Social Anxiety Disorder: A Randomized Controlled Trial. PLOS ONE. 2012;7(5):e37411.

33. Andersson, G, Paxling, B, Roch N P, et al. Internet-Based Psychodynamic versus Cognitive Behavioral Guided Self-Help for Generalized Anxiety Disorder: A Randomized Controlled Trial. Psychotherapy and Psychosomatics. 2012;81(6):344–55.

34. Bani M, Eslam, Ahmad Muayyad. Virtual reality as a distraction technique for pain and anxiety among patients with breast cancer: A randomized control trial. Palliative and Supportive Care. 2019;17(1):29–34.

35. Bell, Caroline J, Colhoun, Helen C, Carter, Frances A, et al. Effectiveness of computerised cognitive behaviour therapy for anxiety disorders in secondary care. 2012;46(7):630–40.

36. Berger, T, Hammerli, K, Gubser, N, et al. Internet-based treatment of depression: a randomized controlled trial comparing guided with unguided self-help. Cogn Behav Ther. 2011;40(4):251–66.

37. Bergström, Jan, Andersson, Gerhard, Ljótsson, Brjánn, et al. Internet-versus group-administered cognitive behaviour therapy for panic disorder in a psychiatric setting: a randomised trial. BMC Psychiatry. 2010;10(1):54.

38. Boele, Florien W, Klein, Martin, Verdonck-de L, Irma M, et al. Internet-based guided self-help for glioma patients with depressive symptoms: a randomized controlled trial. Journal of NeuroOncology. 2018;137(1):191–203.

39. Bond, Gail E, Burr Robert L, Wolf Fredric M, et al. The Effects of a Web-Based Intervention on Psychosocial Well-Being Among Adults Aged 60 and Older With Diabetes. 2010;36(3):446–56.

40. Bowler, Jennifer O, Mackintosh, Bundy, Dunn Barnaby D, et al. A comparison of cognitive bias modification for interpretation and computerized cognitive behavior therapy: Effects on anxiety, depression, attentional control, and interpretive bias. Journal of Consulting and Clinical Psychology. 2012;80(6):1021–33.

41. Braamse, A. M J, van M, B, Visser, O J, et al. A randomized clinical trial on the effectiveness of an intervention to treat psychological distress and improve quality of life after autologous stem cell transplantation. Ann Hematol. 2016;95(1):105–14.

42. Bromberg, J, Wood, M E, Black, R A, et al. A randomized trial of a web-based intervention to improve migraine self-management and coping. Headache. 2012;52(2):244–61.

43. Buhrman, M, Fredriksson, A, Edstrom, G, et al. Guided Internet-delivered cognitive behavioural therapy for chronic pain patients who have residual symptoms after rehabilitation treatment: randomized controlled trial. Eur J Pain. 2013;17(5):753–65.

44. Buhrman, M, Nilsson I E, Jannert, M, et al. Guided internet-based cognitive behavioural treatment for chronic back pain reduces pain catastrophizing: a randomized controlled trial. Journal of rehabilitation medicine. 2011;43(6):500–5.

45. Buhrman, Monica, Skoglund, Astrid, Husell, Josefin, et al. Guided internet-delivered acceptance and commitment therapy for chronic pain patients: A randomized controlled trial. Behaviour Research and Therapy. 2013;51(6):307–15.

46. Buhrman, Monica, Syk, Martin, Burvall, Olle, et al. Individualized Guided Internet-delivered Cognitive-Behavior Therapy for Chronic Pain Patients With Comorbid Depression and Anxiety: A Randomized Controlled Trial. 2015;31(6):504–16.

47. Carlbring, P, Maurin, L, Torngren, C, et al. Individually-tailored, Internet-based treatment for anxiety disorders: A randomized controlled trial. Behav Res Ther. 2011;49(1):18–24.

48. Carrard, I, Crepin, C, Rouget, P, et al. Randomised controlled trial of a guided self-help treatment on the Internet for binge eating disorder. Behav Res Ther. 2011;49(8):482–91.

49. Cohn, M A, Pietrucha, M E, Saslow, L R, et al. An online positive affect skills intervention reduces depression in adults with type 2 diabetes. J Posit Psychol. 2014;9(6):523–34.

50. Cooper, C L, Hind, D, Parry, G D, et al. Computerised cognitive behavioural therapy for the treatment of depression in people with multiple sclerosis: external pilot trial. Trials. 2011;12:259.

51. Damholdt, M F, Mehlsen, M, O’Toole, M S, et al. Web-based cognitive training for breast cancer survivors with cognitive complaints-a randomized controlled trial. Psychooncology. 2016;25(11):1293–300.

52. Dear, Blake F, Gandy, Milena, Karin, Eyal, et al. The Pain Course: a randomised controlled trial examining an internet-delivered pain management program when provided with different levels of clinician support. 2015;156(10):1920–35.

53. Dear, Blake F, Titov, Nick, Perry Kathryn N, et al. The Pain Course: A randomised controlled trial of a clinician-guided Internet-delivered cognitive behaviour therapy program for managing chronic pain and emotional well-being. PAIN®. 2013;154(6):942–50.

54. Devi, Reena, Powell, John, Singh, Sally. A Web-Based Program Improves Physical Activity Outcomes in a Primary Care Angina Population: Randomized Controlled Trial. J Med Internet Res. 2014;16(9):e186.

55. Drozd, F, Skeie, L G, Kraft, P, et al. A web-based intervention trial for depressive symptoms and subjective well-being in patients with chronic HIV infection. AIDS Care. 2014;26(9):1080–9.

56. Engel, C C, Litz, B, Magruder, K M, et al. Delivery of self training and education for stressful situations (DESTRESS-PC): a randomized trial of nurse assisted online self-management for PTSD in primary care. Gen Hosp Psychiatry. 2015;37(4):323–8.

57. Everitt, Hazel, Moss M, Rona, Sibelli Alice, et al. Management of irritable bowel syndrome in primary care: the results of an exploratory randomised controlled trial of mebeverine, methylcellulose, placebo and a self-management website. BMC Gastroenterology. 2013;13(1):68.

58. Farrer, L, Christensen, H, Griffiths, K M, et al. Internet-based CBT for depression with and without telephone tracking in a national helpline: randomised controlled trial. PLoS One. 2011;6(11):e28099.

59. Friesen, Lindsay N, Hadjistavropoulos Heather D, Schneider Luke H, et al. Examination of an internet-delivered cognitive behavioural pain management course for adults with fibromyalgia: a randomized controlled trial. 2017;158(4):593–604.

60. Glozier, Nicholas, Christensen, Helen, Naismith, Sharon, et al. Internet-Delivered Cognitive Behavioural Therapy for Adults with Mild to Moderate Depression and High Cardiovascular Disease Risks: A Randomised Attention-Controlled Trial. PLOS ONE. 2013;8(3):e59139.

61. Hedborg, Kerstin, Muhr, Carin. Multimodal behavioral treatment of migraine: An Internet-administered, randomized, controlled trial. Upsala Journal of Medical Sciences. 2011;116(3):169–86.

62. Hedman, E, Andersson, G, Lindefors, N, et al. Personality change following internet-based cognitive behavior therapy for severe health anxiety. PLoS One. 2014;9(12):e113871.

63. Hedman, Erik, Andersson, Gerhard, Andersson, Erik, et al. Internet-based cognitive– behavioural therapy for severe health anxiety: randomised controlled trial. British Journal of Psychiatry. 2011;198(3):230–6.

64. Hesser, Hugo, Gustafsson, Tore, Lundén, Charlotte, et al. A randomized controlled trial of internet-delivered cognitive behavior therapy and acceptance and commitment therapy in the treatment of tinnitus. Journal of Consulting and Clinical Psychology. 2012;80(4):649–61.

65. Ivarsson, David, Blom, Marie, Hesser, Hugo, et al. Guided internet-delivered cognitive behavior therapy for post-traumatic stress disorder: A randomized controlled trial. Internet Interventions. 2014;1(1):33–40.

66. Jacobi, C, Volker, U, Trockel, M T, et al. Effects of an Internet-based intervention for subthreshold eating disorders: a randomized controlled trial. Behav Res Ther. 2012;50(2):93–9.

67. Jasper, K, Weise, C, Conrad, I, et al. Internet-based guided self-help versus group cognitive behavioral therapy for chronic tinnitus: a randomized controlled trial. Psychotherapy and psychosomatics. 2014;83(4):234–46.

68. Johansson, Birgitta, Bjuhr, Helena, Karlsson, Magdalena, et al. Mindfulness-Based Stress Reduction (MBSR) Delivered Live on the Internet to Individuals Suffering from Mental Fatigue After an Acquired Brain Injury. Mindfulness. 2015;6(6):1356–65.

69. Johnston, Luke, Titov, Nickolai, Andrews, Gavin, et al. A RCT of a Transdiagnostic Internet-Delivered Treatment for Three Anxiety Disorders: Examination of Support Roles and Disorder-Specific Outcomes. PLOS ONE. 2011;6(11):e28079.

70. Knaevelsrud, C, Brand, J, Lange, A, et al. Web-based psychotherapy for posttraumatic stress disorder in war-traumatized Arab patients: randomized controlled trial. J Med Internet Res. 2015;17(3):e71.

71. Kok, Robin N, van S, Annemieke, Beekman, Aartjan TF, et al. Short-Term Effectiveness of Web-Based Guided Self-Help for Phobic Outpatients: Randomized Controlled Trial. J Med Internet Res. 2014;16(9):e226.

72. Kraaij, Vivian, van E, Arnold, Garnefski, Nadia, et al. Effects of a cognitive behavioral self-help program and a computerized structured writing intervention on depressed mood for HIV-infected people: A pilot randomized controlled trial. Patient Education and Counseling. 2010;80(2):200–4.

73. Kuhn, Eric, Kanuri, Nitya, Hoffman Julia E, et al. A randomized controlled trial of a smartphone app for posttraumatic stress disorder symptoms. Journal of Consulting and Clinical Psychology. 2017;85(3):267–73.

74. Lewis, C E, Farewell, D, Groves, V, et al. Internet-based guided self-help for posttraumatic stress disorder (PTSD): Randomized controlled trial. Depress Anxiety. 2017;34(6):555–65.

75. Littleton, H, Grills, A E, Kline, K D, et al. The From Survivor to Thriver program: RCT of an online therapist-facilitated program for rape-related PTSD. J Anxiety Disord. 2016;43:41–51.

76. Ljótsson, B, Hedman, E, Andersson, E, et al. Internet-delivered exposure-based treatment vs. stress management for irritable bowel syndrome: a randomized trial. The American journal of gastroenterology. 2011;106(8):1481–91.

77. Ljótsson Brjánn, Falk, Lisa, Vesterlund, Amanda W, et al. Internet-delivered exposure and mindfulness based therapy for irritable bowel syndrome – A randomized controlled trial. Behaviour Research and Therapy. 2010;48(6):531–9.

78. Lundgren, Johan G, Dahlström Örjan, Andersson Gerhard, et al. The Effect of Guided Web-Based Cognitive Behavioral Therapy on Patients With Depressive Symptoms and Heart Failure: A Pilot Randomized Controlled Trial. J Med Internet Res. 2016;18(8):e194.

79. Mailey, Emily L, Wójcicki Thomas R, Motl Robert W, et al. Internet-delivered physical activity intervention for college students with mental health disorders: A randomized pilot trial. Psychology, Health & Medicine. 2010;15(6):646–59.

80. Migliorini, C, Sinclair, A, Brown, D, et al. A randomised control trial of an Internet-based cognitive behaviour treatment for mood disorder in adults with chronic spinal cord injury. Spinal Cord. 2016;54(9):695–701.

81. Newby, J M, Mackenzie, A, Williams, A D, et al. Internet cognitive behavioural therapy for mixed anxiety and depression: a randomized controlled trial and evidence of effectiveness in primary care. Psychol Med. 2013;43(12):2635–48.

82. Newby, Jill M, Williams Alishia D, Andrews Gavin. Reductions in negative repetitive thinking and metacognitive beliefs during transdiagnostic internet cognitive behavioural therapy (iCBT) for mixed anxiety and depression. Behaviour Research and Therapy. 2014;59:52–60.

83. Newby, Jill, Robins, Lisa, Wilhelm, Kay, et al. Web-Based Cognitive Behavior Therapy for Depression in People With Diabetes Mellitus: A Randomized Controlled Trial. J Med Internet Res. 2017;19(5):e157.

84. Nordgren, Lise B, Hedman, Erik, Etienne, Julie, et al. Effectiveness and cost-effectiveness of individually tailored Internet-delivered cognitive behavior therapy for anxiety disorders in a primary care population: A randomized controlled trial. Behaviour Research and Therapy. 2014;59:1–11.

85. Paxling, B, Almlov, J, Dahlin, M, et al. Guided internet-delivered cognitive behavior therapy for generalized anxiety disorder: a randomized controlled trial. Cogn Behav Ther. 2011;40(3):159–73.

86. Peters, Madelon L, Smeets, Elke, Feijge, Marion, et al. Happy Despite Pain: A Randomized Controlled Trial of an 8-Week Internet-delivered Positive Psychology Intervention for Enhancing Well-being in Patients With Chronic Pain. The Clinical journal of pain. 2017;33(11):962–75.

87. Possemato, Kyle, Kuhn, Eric, Johnson, Emily, et al. Using PTSD Coach in primary care with and without clinician support: a pilot randomized controlled trial. General Hospital Psychiatry. 2016;38:94–8.

88. Robinson, Emma, Titov, Nickolai, Andrews, Gavin, et al. Internet Treatment for Generalized Anxiety Disorder: A Randomized Controlled Trial Comparing Clinician vs. Technician Assistance. PLOS ONE. 2010;5(6):e10942.

89. Rosmarin, David H, Pargament Kenneth I, Pirutinsky, Steven, et al. A randomized controlled evaluation of a spiritually integrated treatment for subclinical anxiety in the Jewish community, delivered via the Internet. Journal of Anxiety Disorders. 2010;24(7):799–808.

90. Roy B, Peter, Craske, Michelle G, Sullivan, Greer, et al. Delivery of Evidence-Based Treatment for Multiple Anxiety Disorders in Primary Care: A Randomized Controlled Trial. JAMA. 2010;303(19):1921–8.

91. Ruehlman, L S, Karoly, P, Enders, C. A randomized controlled evaluation of an online chronic pain self management program. Pain. 2012;153(2):319–30.

92. Ruwaard, Jeroen, Broeksteeg, Janneke, Schrieken, Bart, et al. Web-based therapist-assisted cognitive behavioral treatment of panic symptoms: A randomized controlled trial with a three-year follow-up. Journal of Anxiety Disorders. 2010;24(4):387–96.

93. Sanchez O, V C, Munro, C, Stahl, D, et al. A randomized controlled trial of internet-based cognitive-behavioural therapy for bulimia nervosa or related disorders in a student population. Psychol Med. 2011;41(2):407–17.

94. Seekles, Wike, van S, Annemieke, Beekman, Aartjan, et al. Effectiveness of guided self-help for depression and anxiety disorders in primary care: A pragmatic randomized controlled trial. Psychiatry Research. 2011;187(1):113–20.

95. Sexton, Minden B, Byrd Michelle R, O’Donohue William T, et al. Web-based treatment for infertility-related psychological distress. Archives of Women’s Mental Health. 2010;13(4):347–58.

96. Shigaki, Cheryl L, Smarr Karen L, Siva, Chokkalingam, et al. RAHelp: An Online Intervention for Individuals With Rheumatoid Arthritis. 2013;65(10):1573–81.

97. Silfvernagel, Kristin, Carlbring, Per, Kabo, Julia, et al. Individually Tailored Internet-Based Treatment for Young Adults and Adults With Panic Attacks: Randomized Controlled Trial. J Med Internet Res. 2012;14(3):e65.

98. Spence, J, Titov, N, Johnston, L, et al. Internet-based trauma-focused cognitive behavioural therapy for PTSD with and without exposure components: a randomised controlled trial. J Affect Disord. 2014;162:73–80.

99. Spence, Jay, Titov, Nickolai, Dear Blake F, et al. Randomized controlled trial of Internet-delivered cognitive behavioral therapy for posttraumatic stress disorder. 2011;28(7):541–50.

100. Titov, N, Andrews, G, Davies, M, et al. Internet treatment for depression: a randomized controlled trial comparing clinician vs. technician assistance. PLoS One. 2010;5(6):e10939.

101. Titov, Nickolai, Andrews, Gavin, Johnston, Luke, et al. Transdiagnostic Internet treatment for anxiety disorders: A randomized controlled trial. Behaviour Research and Therapy. 2010;48(9):890–9.

102. Trompetter, H R, Bohlmeijer, E T, Veehof, M M, et al. Internet-based guided self-help intervention for chronic pain based on Acceptance and Commitment Therapy: a randomized controlled trial. Journal of behavioral medicine. 2015;38(1):66–80.

103. Trudeau, Kimberlee J, Pujol Lynette A, DasMahapatra, Pronabesh, et al. A randomized controlled trial of an online self-management program for adults with arthritis pain. Journal of Behavioral Medicine. 2015;38(3):483–96.

104. van B, Wouter, Riper, Heleen, Klein, Britt, et al. An Internet-Based Guided Self-Help Intervention for Panic Symptoms: Randomized Controlled Trial. J Med Internet Res. 2013;15(7):e154.

105. Varley, Rachel, Webb Thomas L, Sheeran, Paschal. Making self-help more helpful: A randomized controlled trial of the impact of augmenting self-help materials with implementation intentions on promoting the effective self-management of anxiety symptoms. Journal of Consulting and Clinical Psychology. 2011;79(1):123–8.

106. Vernmark, K, Lenndin, J, Bjarehed, J, et al. Internet administered guided self-help versus individualized e-mail therapy: A randomized trial of two versions of CBT for major depression. Behav Res Ther. 2010;48(5):368–76.

107. Weise, C, Kleinstäuber, M, Andersson, G. Internet-Delivered Cognitive-Behavior Therapy for Tinnitus: A Randomized Controlled Trial. Psychosomatic medicine. 2016;78(4):501–10.

108. Willems, Roy A, Mesters, Ilse, Lechner, Lilian, et al. Long-term effectiveness and moderators of a web-based tailored intervention for cancer survivors on social and emotional functioning, depression, and fatigue: randomized controlled trial. Journal of Cancer Survivorship. 2017;11(6):691–703.

109. Williams, D A, Kuper, D, Segar, M, et al. Internet-enhanced management of fibromyalgia: a randomized controlled trial. Pain. 2010;151(3):694–702.

110. Wilson, M, Roll, J M, Corbett, C, et al. Empowering Patients with Persistent Pain Using an Internet-based Self-Management Program. Pain management nursing : official journal of the American Society of Pain Management Nurses. 2015;16(4):503–14.

111. Wilson, Marian, Hewes, Casey, Barbosa L, Celestina, et al. Engaging Adults With Chronic Disease in Online Depressive Symptom Self-Management. 2017;40(6):834–53.

112. Wims, Edward, Titov, Nickolai, Andrews, Gavin, et al. Clinician-Assisted Internet-Based Treatment is Effective for Panic: A Randomized Controlled Trial. 2010;44(7):599–607.

113. Wootton, Bethany M, Dear Blake F, Johnston, Luke, et al. Remote treatment of obsessive-compulsive disorder: A randomized controlled trial. Journal of Obsessive-Compulsive and Related Disorders. 2013;2(4):375–84.

114. Yun, Y H, Lee, K S, Kim, Y W, et al. Web-based tailored education program for disease-free cancer survivors with cancer-related fatigue: a randomized controlled trial. J Clin Oncol. 2012;30(12):1296–303.

